# Cognition and Electrophysiology Clustering in Clinical High Risk for Psychosis Delineates Distinct Dimensions of Heterogeneity: Implications for Multimodal Clustering

**DOI:** 10.64898/2026.03.14.26347633

**Authors:** Walid Yassin, James B. Green, Michael Cai, Danah Ansari, Xue-Jun Kong, Elisabetta C. del Re, Holly K. Hamilton, Spero Nicholas, Brian J. Roach, Peter M. Bachman, Aysenil Belger, Ricardo E. Carrión, Erica Duncan, Jason K. Johannesen, Gregory A. Light, Sandra K. Loo, Margaret A. Niznikiewicz, Jean Addington, Carrie E. Bearden, Kristin S. Cadenhead, Tyrone D. Cannon, Diana O. Perkins, Elaine F. Walker, Scott W. Woods, Matcheri Keshavan, Daniel H. Mathalon, William S. Stone

## Abstract

Individuals at clinical high risk for psychosis (CHR) are cognitively and neurobiologically heterogeneous, which encourages the use of a clustering approach to parse this heterogeneity. Multimodal approaches are assumed to be superior to unimodal approaches in identifying subgroups. With the success of the use of cognition and electrophysiological measures collectively in established psychotic disorders, and the lack of such an approach in CHR, we were motivated to address this gap. Using the North American Psychosis-Risk Longitudinal Study (NAPLS) 2 consortia (CHR (N=764)), we applied unsupervised cluster analysis on the combined cognitive and electrophysiology measures to identify CHR subgroups and assess their relationship with clinical and functional outcomes. A two-cluster solution with modest separability was found, which prompted the use of an alternative probabilistic, rather than discrete, clustering approach. Individuals who were more likely to be in Cluster 1 exhibited poorer cognitive performance, larger N100, mismatch negativity, and P300 amplitudes, and worse functioning, as well as a younger age of onset. These findings were largely replicated in NAPLS 3 (CHR (N=628)). Taken together, the results of our previous study of cognition-only clustering and the current study of combining cognition and electrophysiology indicate that multimodal clustering, if not developmentally informed, may obscure meaningful subtyping.

## Introduction

Classical diagnostic categories in psychiatry can obscure the underlying biological heterogeneity as well as the dynamic nature of mental health conditions.^1^ This is especially important in the early stages of psychopathology, such as the clinical high risk for psychosis (CHR) syndrome, where detection, risk stratification, and preventive interventions are likely to have their greatest impact. A large proportion of CHR individuals experience cognitive decline, information processing challenges, persistent functional impairment, and other clinical and non-clinical symptoms.^1–3^

Cognitive impairment in the psychosis spectrum, including CHR, is well documented.^1,3,4^ Deficits in several cognitive domains are present much earlier than illness onset and are typically associated with impaired functioning and increased risk for transition to full psychosis.^1,4,5^ Importantly, these deficits tend to be stable over time and are associated with genetic liability for schizophrenia, which reflects their trait-like nature and positions them as plausible markers of neurodevelopmental vulnerability.^1,5–9^

Information processing aberrations, indexed via electroencephalography (EEG)-based assessments, are also present in the CHR population.^2,10,11^ A large body of evidence demonstrates amplitude reduction of early auditory sensory event-related potential (ERP) components, including mismatch negativity (MMN) and N100 (N1), in schizophrenia versus those who are typically developing, which has also been demonstrated, to a lesser extent, in CHR.^10–26^ This has been linked to challenges in predictive coding and sensory integration, among others.^17,24,27–29^ Reduced and delayed P300 ERP responses to target and/or novel stimuli in oddball tasks are also reported in the psychosis spectrum, and related to attentional resource allocation as well as internal model updating.^2,14,22,23,23,30–38^

Individuals at the CHR stage are at increased risk for transition to a psychotic disorder, most often schizophrenia.^39^ However, the path to transition is neither linear nor deterministic. Most individuals at CHR do not transition; only about 17-30% do, after a period of 2-3 years from their initial CHR diagnosis.^40,41^ As such, a large body of evidence demonstrates that CHR is not a static and uniform condition, but rather a biologically heterogeneous and dynamic one.^42^ In alignment with the current biological evidence and biomarker-informed approaches in psychiatry, data-driven machine learning techniques have been increasingly used to parse heterogeneity in the CHR population.^1^ Such approaches have been used to inform prognosis, treatment selection, and mechanistic understanding of the condition.^43^

Several of our studies and those from other groups have attempted to better understand the heterogeneity and dynamic nature of psychosis, tackling its different stages.^1,9^ The Bipolar-Schizophrenia Network on Intermediate Phenotypes group, for example, focused on established stages of the psychosis spectrum, including schizophrenia, bipolar disorder with psychosis, and schizoaffective disorder. In one of their studies, the group applied a clustering technique on cognition and electrophysiology variables, which resulted in three clusters or “Biotypes”; biotype 1 was characterized by reduced neural activity and severe cognitive impairment, biotype 2 by increased neural activity and moderate to severe cognitive impairment, and biotype 3 was similar to controls.^44^ Another study by Lee et al, using cognition and electrophysiology data from bipolar, schizophrenia, and control subjects, found a two-cluster solution, “intact” and “impaired”.^45^ In antipsychotic naive first episode patients, Bak et al. used a Gaussian Mixture Model on cognition and electrophysiology data and found two main subgroups.^46^ Collectively, this work provided a better understanding of the underlying biological overlap and heterogeneity of psychotic disorders. Thus, we hypothesize that multimodal clustering, using cognition and electrophysiology, would yield separable subgroups in CHR.

To our knowledge, no other published studies have yet investigated the potential CHR clusters resulting from both cognition and electrophysiology modalities jointly. The North American Psychosis-Risk Longitudinal Studies (NAPLS) provide a unique opportunity to evaluate this in large, deeply characterized CHR cohorts. Using both NAPLS-2 and NAPLS-3, the current study utilizes a clustering approach on combined cognitive and electrophysiological measures to identify subgroups within CHR individuals and evaluate them on functional and clinical measures.

## Methods

### Participants

The participants’ data were obtained from the NAPLS-2 (CHR (n=764) HC (n=280)) and NAPLS-3 (CHR (n=628) HC (n=84)).^47,48^ The CHR participants met the Criteria of Psychosis-Risk Syndromes (COPS).^49^ The clinical vignettes for each CHR participant were reviewed to obtain a consensus diagnosis after a comprehensive assessment, which included administering the Structured Interview for Pyschosis-Risk Syndromes (SIPS) in both studies, the Structured Clinical Interview for DSM (SCID) IV^50^ for NAPLS-2 and the SCID-5 for NAPLS-3.^47–49^ All sites (Emory University, Harvard University, University of Calgary, University of California, Los Angeles, University of California San Francisco, University of California San Diego, University of North Carolina at Chapel Hill, Yale University, and Zucker Hillside Hospital) provided Institutional Review Board approval. After receiving a complete description of the study, adult participants signed an informed consent, whereas participants who are minors provided written assent, with parents/guardians signing informed consent.

Inclusion and exclusion criteria: Participants’ ages ranged between 12 and 35 years old and were excluded if they had an IQ less than 70, a history of a central nervous system disorder, apparent psychosis risk symptoms better explained by another Axis I disorder, or a current or lifetime Axis I psychotic disorder. If there was retrospective evidence that an individual only met CHR criteria and was not fully psychotic when anti-psychotic medication was started, prior or current antipsychotic treatment at study entry was not exclusionary. None of the following criteria were met for healthy controls (HC): i) any psychosis risk syndrome, ii) current or past psychotic disorder, or iii) Cluster A personality disorder diagnosis. Potential HC who were currently using psychotropic medication were excluded. Those having first-degree relatives with a history of a psychotic disorder or other disorders involving psychotic symptoms were also excluded. Details on the diagnosis, inclusion, and exclusion criteria can be found elsewhere.^47,48^

### Assessments

#### Neurocognition

Neurocognitive measures from the MATRICS Consensus Cognitive Battery (MCCB) were utilized in both NAPLS-2 and-3 consortia, including the Wide Range Achievement Test, Fourth Edition (WRAT-4)^51^, Wechsler Abbreviated Scale of Intelligence II (WASI-II) IQ and Vocabulary^52^, Hopkins Verbal Learning Test-Revised (HVLT-R)^53^, Brief Assessment of Cognition in Schizophrenia Symbol Coding (BACS-SC)^54^, Letter-Number Span (LNS)^55^, and A-Continuous Performance Test (A-CPT), which comprised vigilance (QA-*Vigilance*) and high working memory load/no interference (Q3A-MEM) conditions^56^ (See Yassin et al. 2025). The A-CPT interference condition, Q3INT (the letter “A” comes four letters after “Q”), was used in NAPLS-2, and the Q1AINT (where the letter “A” comes two letters after “Q”) was used in NAPLS-3.

#### Electrophysiology

NAPLS-2 baseline EEG data were collected at all sites using either 32- or 64-channel BioSemi ActiveTwo recording systems. Paradigms included a mismatch negativity paradigm with pitch, duration, and pitch+duration double-deviants, which were delivered concurrently with a visual oddball task (for details, see Hamilton et al.^11,35^). The visual oddball paradigm involved frequent standard stimuli (small blue circle), infrequent target stimuli (large blue circle) requiring a button press response used to elicit the target P300 (P3b), and infrequent novel non-target fractal images used to elicit novelty P300 (P3a).^35^ A separate 3-stimulus auditory oddball paradigm was used to elicit auditory P3b to target tones and P3a to novel sounds.^2^ N100 to auditory stimuli were also assessed (see Duncan et al.^10^).

NAPLS-3 baseline EEG data were collected at all sites using a 64-channel BioSemi Active Two system. EEG data from NAPLS-3 included in the current analysis were derived from a 16-minute set of paradigms shared with other consortia (PSYSCAN and PRONIA) to harmonize a set of EEG-based measures as part of an NIH initiative known as HARMONY.^57–59^ The HARMONY session comprised a single long-duration deviant MMN paradigm that ran concurrently with a 2-stimulus visual oddball task. The visual oddball task included frequent standards (small blue circle) and infrequent target stimuli (large blue circle) used to elicit target P3b. A separate 2-stimulus auditory oddball task was used to elicit auditory target P3b, with tones identical to those used in NAPLS-2. Auditory N100 was assessed in response to auditory standards and targets. For general details about the NAPLS3 site recording procedures, see Jacob et al (2021).^60^

#### Clinical and Functional Measures

Clinical and functional measures included in this study were the Scale of Psychosis-Risk Symptoms (SOPS), Global Assessment of Functioning (GAF),^61^ and the Global Functioning Social (GF: Social) and Role Scales (GF: Role).^62^ The SIPS Presence of Psychotic Symptoms criteria were used to determine the current clinical status, including conversion to psychosis, persistence/progression of CHR symptoms, or remission from the CHR syndrome, at each clinical assessment over the two-year follow-up period.

### Machine learning

#### Preprocessing

NAPLS-2 and-3 were similarly processed. Instances, or participants with more than 50% missingness in the input variables, were excluded. NAPLS-2 was the discovery sample, and NAPLS-3 was the external validation sample. The processing included imputation of missing data, site correction, removal of typically developing age and sex effects from HC, data standardization, and principal component analysis (PCA). Principal components (PCs)s accounting for a cumulative variance of 80% were used. The variable loadings for each PC were produced at this step. A detailed description can be found elsewhere.^1^

#### Clustering algorithm

The clustering was performed on the combined cognition and electrophysiology data using the R package clValid v0.7.^63^ DIvisive ANAlysis Clustering (DIANA) was used to cluster the data.^63^ DIANA was found to be an optimum classifier in both NAPLS-2 and-3 in our previous work.^1^ K-means and AGglomerative NESting (Agnes) were also tested in the case that those algorithms would perform better on this specific dataset distribution, including the combined cognition and EEG data.

*Post hoc* model-based clustering was used to extract measures of uncertainty. Specifically, Gaussian Mixture Modeling (GMM) was performed because it provides an established method of extracting the membership uncertainty. The mclust package (Scrucca et al., 2016) in R was utilized for model-based clustering. The Bayesian Information Criterion (BIC) was computed for a range of models using the mclustBIC() function to guide model selection. The selected model was fitted with the Expectation-Maximization algorithm. The resulting model provided the number of clusters, the model type (covariance structure), and posterior probabilities of class membership for each observation. Finally, the posterior probabilities were extracted and used for further analysis. This step was performed for NAPLS-2 and-3. The posterior probability described the probability of a participant belonging to cluster 1 (so, values closer to 0 indicated a stronger likelihood for cluster 2 membership).

Clustering was also performed *post hoc* on the EEG-only data using DIANA, Agnes, and K-means to better explain the results of the combined EEG and cognition analysis.

## Statistical Analysis

Demographic summary statistics were reported for the CHR clusters in both consortia, and associations between demographics and posterior probabilities were assessed. Age was included as a covariate in subsequent analyses, given its significant correlation with posterior probabilities. Since “years of education”, which was also significantly correlated with posterior probabilities, had no impact on the results when performing sensitivity analyses with and without it, only age was included in subsequent analyses to simplify the model.

To delineate the characteristics of the clusters’ membership probabilities, associations between cognitive and EEG variables and posterior probabilities were evaluated. Since the variables did not meet the normality assumption, Spearman’s rank-order correlations were implemented. Next, Spearman’s rank order correlations analyzed the association of posterior probabilities with clinical measures (including age of onset) and functional outcomes. Unadjusted and partial correlations adjusting for age are shown, given that age was significantly associated with posterior probabilities. False Discovery Rate p-value corrections were used within clinical and functional analyses. Differences in posterior probabilities across clinical outcomes, including converter, remitter, and non-remitter (i.e., persistent CHR) status at the last follow-up assessment, were assessed with and without controlling for age using ANOVA and ANCOVA, respectively.

All analytic steps were conducted in NAPLS-2 as the main analyses, and then in NAPLS-3 for external validation purposes. The reported posterior probabilities are the probabilities of a participant being in cluster 1, so positive correlations with a variable indicate that higher values of that variable are associated with a greater likelihood of cluster 1 membership, while negative correlations suggest association of higher values with greater likelihood of cluster 2 membership.

Statistical analyses were conducted using R version v4.2.

## Results

NAPLS-2 and NAPLS-3 demographic data are presented in Table S1.

### Clustering

Using the combined cognitive and EEG data from NAPLS-2, the algorithm having the highest silhouette score of.30 was DIANA for two clusters. NAPLS-3 clustering yielded similar results with a highest silhouette score of 0.40 using DIANA for a two-cluster solution. Silhouette scores, which range from-1 to 1, used to identify the optimal clustering algorithm, are presented in Supplementary Table S2. Using the combined cognitive and EEG data from NAPLS-2, the optimal cluster number with the highest BIC value (BIC=-19,329.61) was two clusters via the VEI model (Diagonal distribution, variable volume, and equal shape).^64^ Using GMM on the NAPLS-3 data also resulted in two clusters using the VEI model (BIC=-20,222.30). The *post hoc* results of the NAPLS-2 EEG only clustering demonstrated the highest silhouette score of 0.34 using DIANA (Table S2).

### NAPLS-2

#### Demographics

CHR participants who are more likely to be in cluster 2 (C2) were relatively older (p < 0.001) and had higher educational attainment than participants likely to be in cluster 1 (C1). No differences in sex or household income between the C1 and C2 clusters were identified (Table 1).

**Table 1:** Cluster comparison of CHR participants’ demographic data (NAPLS2).

| | Cluster 1 (N=310) | | Cluster 2 (N=275) | | $\rho/F$ | DF | P |
| --- | --- | --- | --- | --- | --- | --- | --- |
|  | M/N | SD/% | M/N | SD/% |  |  |  |
| Age | 18.07 | 4.52 | 19.06 | 4.10 | -0.2 | – | <0.001*** |
| Education (years) | 10.7 | 2.8 | 11.84 | 2.74 | -0.25 | – | <0.001*** |
| Sex (male) | 169 | 54.52% | 169 | 61.45% | 2.44 | 1 | 0.12 |
| Household income |  |  |  |  | 1.92 | 5 | 0.09 |
| Less than 10,000 | 229 | 80.35% | 197 | 75.77% |  |  |  |
| 10,000 – 19,999 | 25 | 8.77% | 36 | 13.85% |  |  |  |
| 20,000 – 39,999 | 17 | 5.96% | 17 | 6.54% |  |  |  |
| 40,000 – 59,999 | 8 | 2.81% | 4 | 1.54% |  |  |  |
| 60,000 – 99,999 | 5 | 1.75% | 3 | 1.15% |  |  |  |
| 100,000 + | 1 | 0.35% | 3 | 1.15% |  |  |  |

CHR: Clinical high risk for psychosis; M/N: Mean / Category count; SD/%: Standard deviation/ Category percentage; $\rho/F$ : Overall Spearman-rank correlation coefficient/ ANOVA F-statistics; DF: Degrees of freedom; P: P-value; posterior probabilities are the probabilities of a participant being in cluster 1.

#### Cognition and EEG

Despite partial overlap, cluster membership demonstrated significant associations with both cognitive and EEG data (Table 2, Figures 1, 2). Cognition was higher across the board, with a small to moderate effect size in C2 compared to C1. The EEG data showed C1, relative to C2, membership to be associated with larger ERP component amplitudes, including more negative N1 (rho= |.291∼.455|) and MMN (rho=|.205∼.233|) and more positive P3 (rho= |.087∼.2|) at parietal (P3, P4, and Pz; auditory and visual), central ( C3, C4, and Cz; only visual), and frontal ((F3, F4, and Fz; only visual) electrode sites. Visual P3 latency was also earlier for the C1, relative to the C2 cluster.

**Figure 1:**
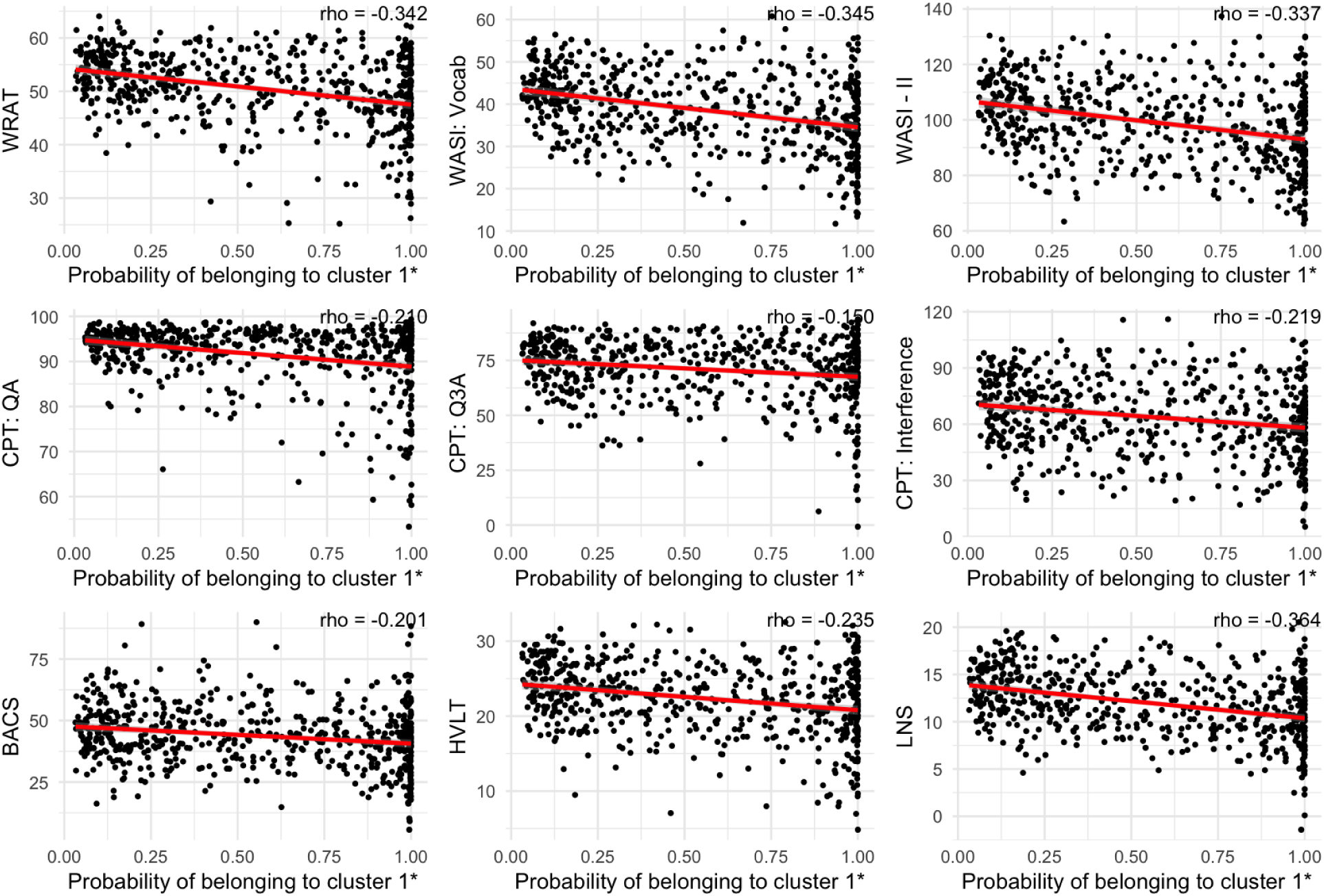
Correlations between cognitive variables with posterior probabilities in CHR participants from NAPLS2. *CHR: Clinical high risk for psychosis; WRAT: Wide Range Achievement Test-Four Reading subtest; WASI Vocab: Wechsler Abbreviated Scale for Intelligence-2 Vocabulary; WASI - II: Weschler Abbreviated Scale for Intelligence - 2: Intelligence Quotient; CPT: Auditory Working Memory Continuous Performance Test; QA: Vigilance; Q3A: Working Memory Load/No Interference; BACS: Brief Assessment of Cognition in Schizophrenia – Symbol Coding; HVLT: Hopkins Verbal Learning Test-Revised; LNS: Letter-Number-Span; *: p<0.05; probabilities of a participant being in cluster 1 are posterior probabilities. Cognition variables plotted with the probability of belonging to cluster 1*.

**Figure 2.**
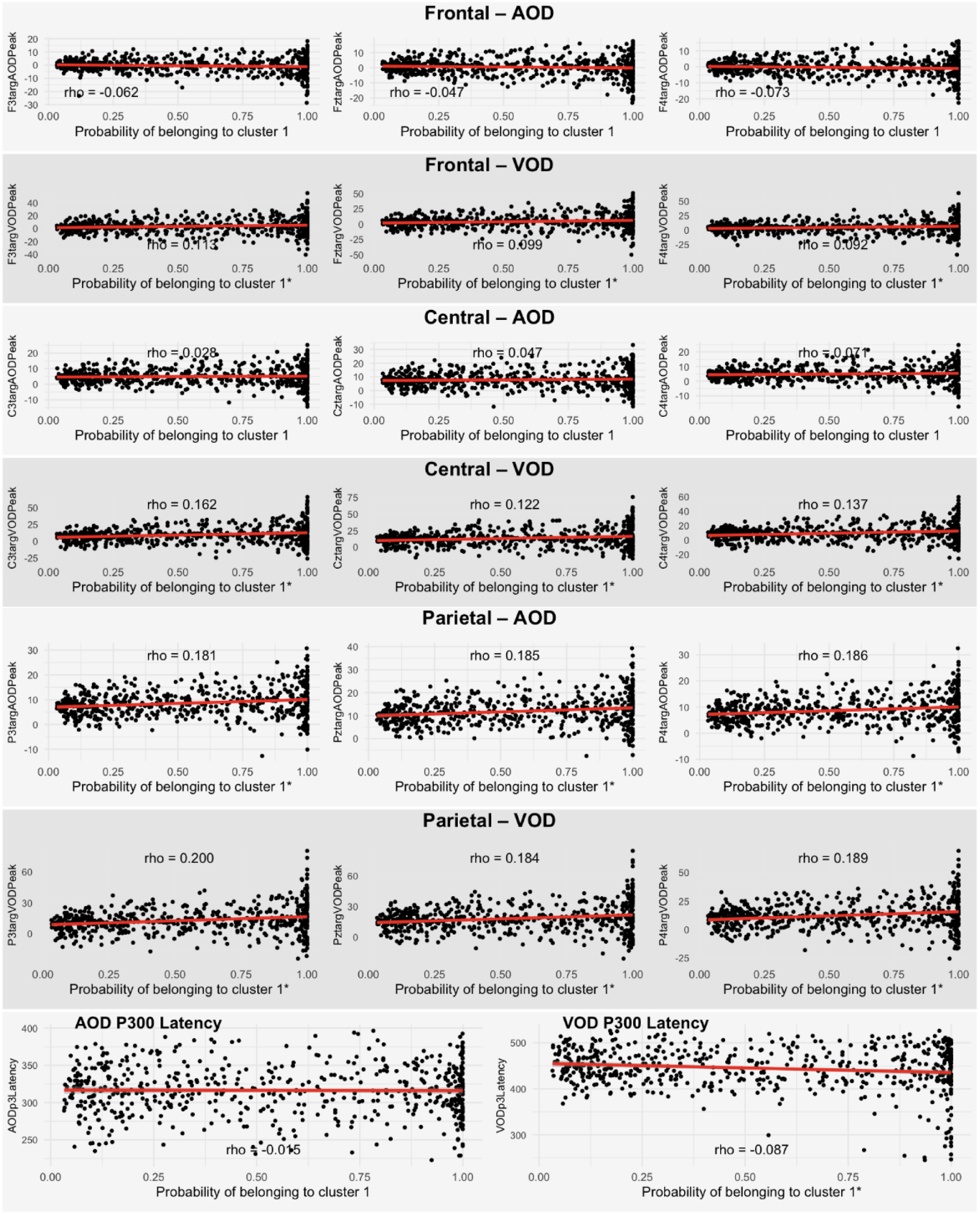
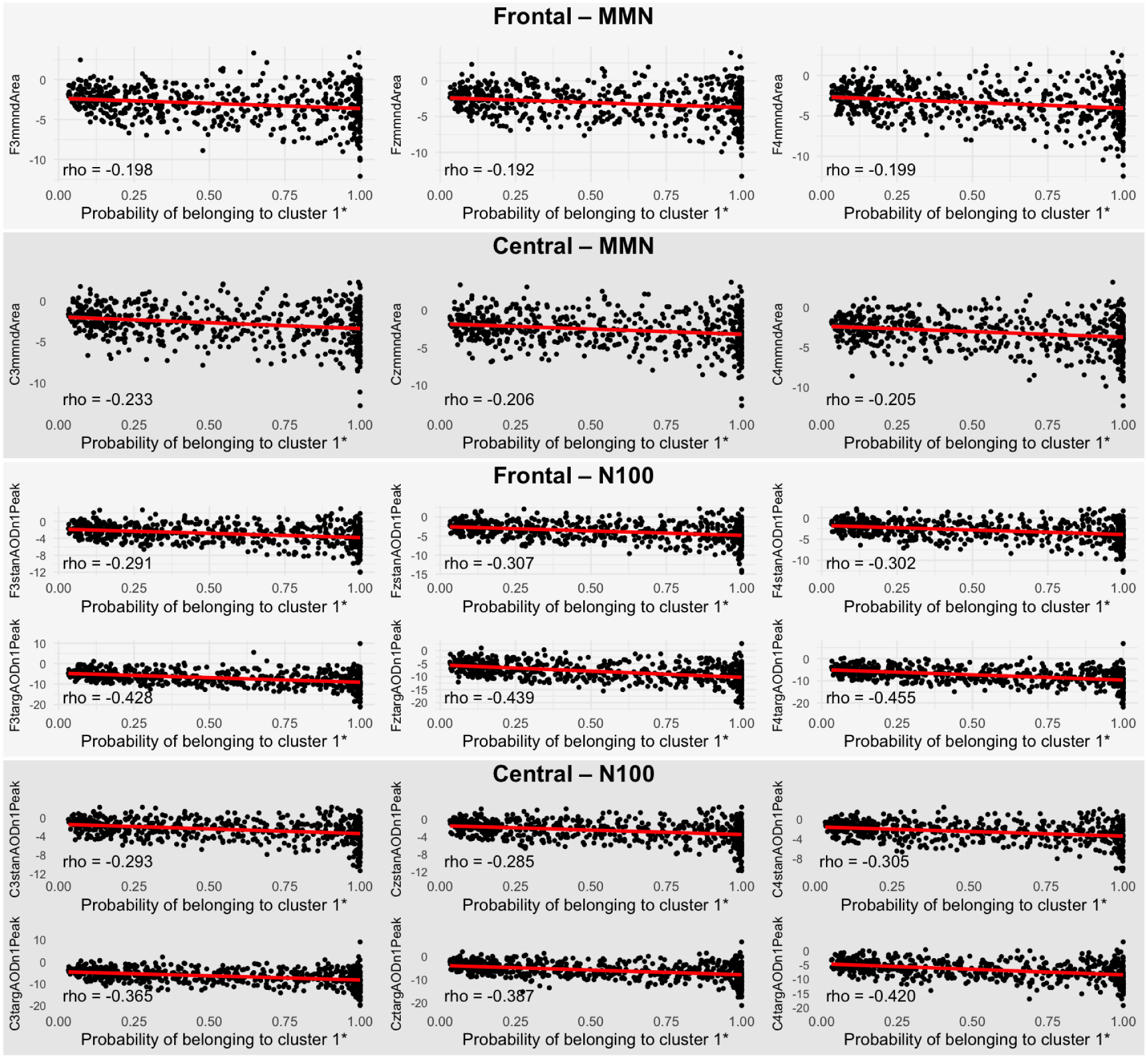
Correlations between electrophysiology variables with posterior probabilities in CHR participants from NAPLS2. *CHR: Clinical high risk for psychosis; MMN: Missmatch negativity; AOD: Auditory Oddball task; VOD: Visual Oddball Task; N1 (Or N100): Event related potential Negative peak around 100 milliseconds; P3 (Or P300): Event related potential Positive peak around 300 milliseconds; Targ: Target; Stan: Standard; *: p<0.05; probabilities of a participant being in cluster 1 are posterior probabilities*

**Table 2:**
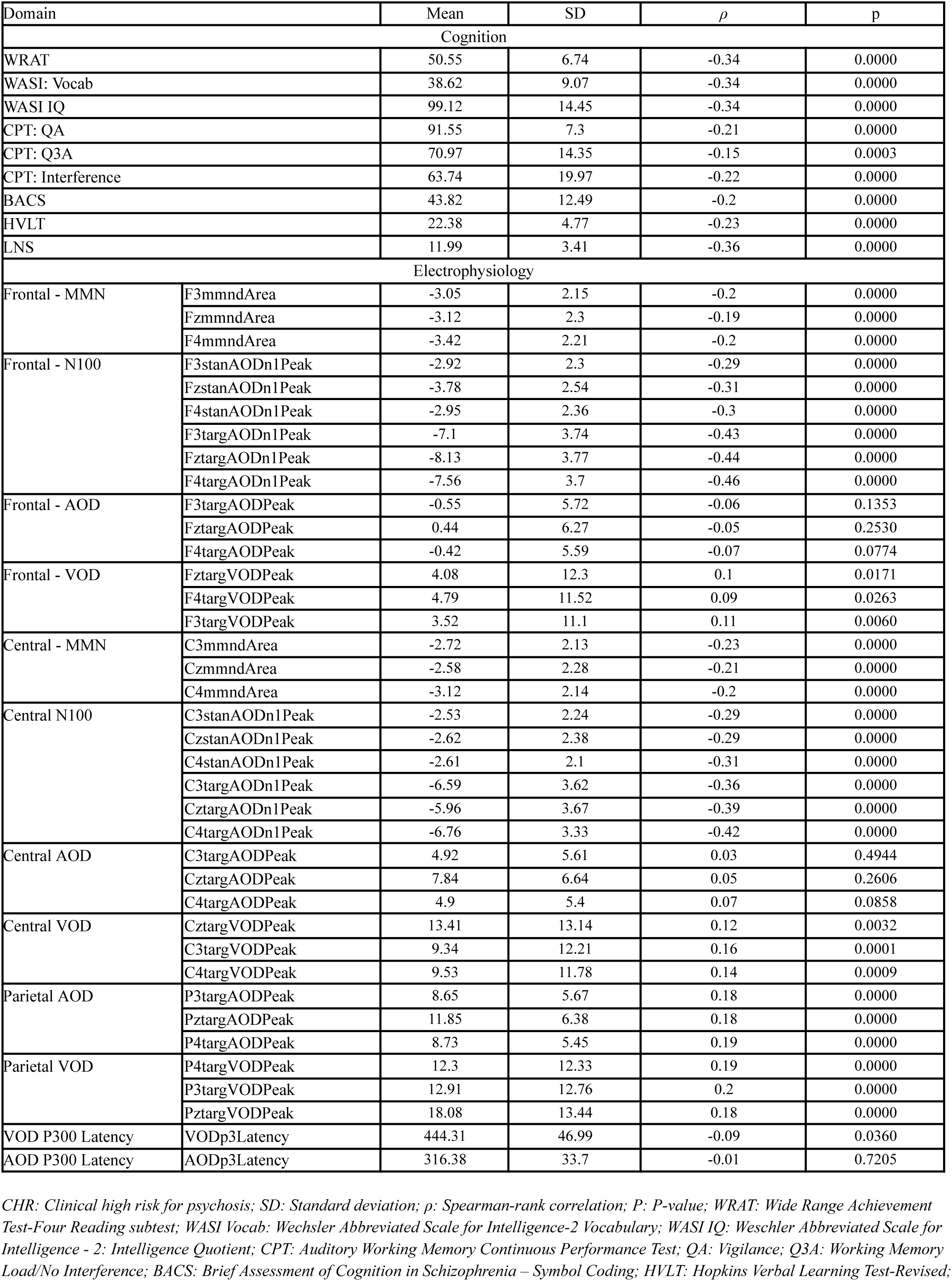
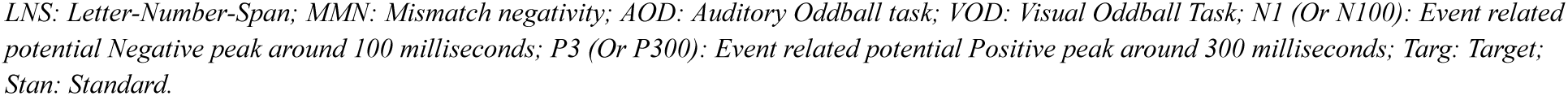
Correlations between cognitive and electrophysiology variables with posterior probabilities in CHR participants from NAPLS2.

| Domain | | Mean | SD | $\rho$ | p |
| --- | --- | --- | --- | --- | --- |
| Cognition |  |  |  |  |  |
| WRAT |  | 50.55 | 6.74 | -0.34 | 0.0000 |
| WASI: Vocab |  | 38.62 | 9.07 | -0.34 | 0.0000 |
| WASI IQ |  | 99.12 | 14.45 | -0.34 | 0.0000 |
| CPT: QA |  | 91.55 | 7.3 | -0.21 | 0.0000 |
| CPT: Q3A |  | 70.97 | 14.35 | -0.15 | 0.0003 |
| CPT: Interference |  | 63.74 | 19.97 | -0.22 | 0.0000 |
| BACS |  | 43.82 | 12.49 | -0.2 | 0.0000 |
| HVL |  | 22.38 | 4.77 | -0.23 | 0.0000 |
| LNS |  | 11.99 | 3.41 | -0.36 | 0.0000 |
| Electrophysiology |  |  |  |  |  |
| Frontal - MMN | F3mmndArea | -3.05 | 2.15 | -0.2 | 0.0000 |
|  | FzmmndArea | -3.12 | 2.3 | -0.19 | 0.0000 |
|  | F4mmndArea | -3.42 | 2.21 | -0.2 | 0.0000 |
| Frontal - N100 | F3stanAODn1Peak | -2.92 | 2.3 | -0.29 | 0.0000 |
|  | FzstanAODn1Peak | -3.78 | 2.54 | -0.31 | 0.0000 |
|  | F4stanAODn1Peak | -2.95 | 2.36 | -0.3 | 0.0000 |
|  | F3targAODn1Peak | -7.1 | 3.74 | -0.43 | 0.0000 |
|  | FztargAODn1Peak | -8.13 | 3.77 | -0.44 | 0.0000 |
|  | F4targAODn1Peak | -7.56 | 3.7 | -0.46 | 0.0000 |
| Frontal - AOD | F3targAODPeak | -0.55 | 5.72 | -0.06 | 0.1353 |
|  | FztargAODPeak | 0.44 | 6.27 | -0.05 | 0.2530 |
|  | F4targAODPeak | -0.42 | 5.59 | -0.07 | 0.0774 |
| Frontal - VOD | FztargVODPeak | 4.08 | 12.3 | 0.1 | 0.0171 |
|  | F4targVODPeak | 4.79 | 11.52 | 0.09 | 0.0263 |
|  | F3targVODPeak | 3.52 | 11.1 | 0.11 | 0.0060 |
| Central - MMN | C3mmndArea | -2.72 | 2.13 | -0.23 | 0.0000 |
|  | CzmmndArea | -2.58 | 2.28 | -0.21 | 0.0000 |
|  | C4mmndArea | -3.12 | 2.14 | -0.2 | 0.0000 |
| Central N100 | C3stanAODn1Peak | -2.53 | 2.24 | -0.29 | 0.0000 |
|  | CzstanAODn1Peak | -2.62 | 2.38 | -0.29 | 0.0000 |
|  | C4stanAODn1Peak | -2.61 | 2.1 | -0.31 | 0.0000 |
|  | C3targAODn1Peak | -6.59 | 3.62 | -0.36 | 0.0000 |
|  | CztargAODn1Peak | -5.96 | 3.67 | -0.39 | 0.0000 |
|  | C4targAODn1Peak | -6.76 | 3.33 | -0.42 | 0.0000 |
| Central AOD | C3targAODPeak | 4.92 | 5.61 | 0.03 | 0.4944 |
|  | CztargAODPeak | 7.84 | 6.64 | 0.05 | 0.2606 |
|  | C4targAODPeak | 4.9 | 5.4 | 0.07 | 0.0858 |
| Central VOD | CztargVODPeak | 13.41 | 13.14 | 0.12 | 0.0032 |
|  | C3targVODPeak | 9.34 | 12.21 | 0.16 | 0.0001 |
|  | C4targVODPeak | 9.53 | 11.78 | 0.14 | 0.0009 |
| Parietal AOD | P3targAODPeak | 8.65 | 5.67 | 0.18 | 0.0000 |
|  | PztargAODPeak | 11.85 | 6.38 | 0.18 | 0.0000 |
|  | P4targAODPeak | 8.73 | 5.45 | 0.19 | 0.0000 |
| Parietal VOD | P4targVODPeak | 12.3 | 12.33 | 0.19 | 0.0000 |
|  | P3targVODPeak | 12.91 | 12.76 | 0.2 | 0.0000 |
|  | PztargVODPeak | 18.08 | 13.44 | 0.18 | 0.0000 |
| VOD P300 Latency | VODp3Latency | 444.31 | 46.99 | -0.09 | 0.0360 |
| AOD P300 Latency | AODp3Latency | 316.38 | 33.7 | -0.01 | 0.7205 |

CHR: Clinical high risk for psychosis; SD: Standard deviation; $\rho$ : Spearman-rank correlation; P: P-value; WRAT: Wide Range Achievement Test-Four Reading subtest; WASI Vocab: Wechsler Abbreviated Scale for Intelligence-2 Vocabulary; WASI IQ: Wechsler Abbreviated Scale for Intelligence - 2: Intelligence Quotient; CPT: Auditory Working Memory Continuous Performance Test; QA: Vigilance; Q3A: Working Memory Load/No Interference; BACS: Brief Assessment of Cognition in Schizophrenia – Symbol Coding; HVL: Hopkins Verbal Learning Test-Revised;
LNS: Letter-Number-Span; MMN: Mismatch negativity; AOD: Auditory Oddball task; VOD: Visual Oddball Task; N1 (Or N100): Event related potential Negative peak around 100 milliseconds; P3 (Or P300): Event related potential Positive peak around 300 milliseconds; Targ: Target; Stan: Standard.

#### Clinical and functional outcomes

CHR participants who were more likely to be in C2 had significantly higher social functioning (q <0.01) while adjusting for the effects of age (Table 3 & Figures 3). Age at CHR symptom onset was earlier for participants who were more likely to be in C1 than in C2 (q <0.001). No other significant associations between cluster membership and clinical or functional outcomes were identified (Table 3 & Figures 3a-b). Sensitivity analysis demonstrated no differences in outcomes when adding education to the models (Table S3). Pearson’s correlations between duration of illness and uncertainty demonstrated a negative relationship (r:-0.10, p= 0.01) (Figure S1).

**Figure 3.**
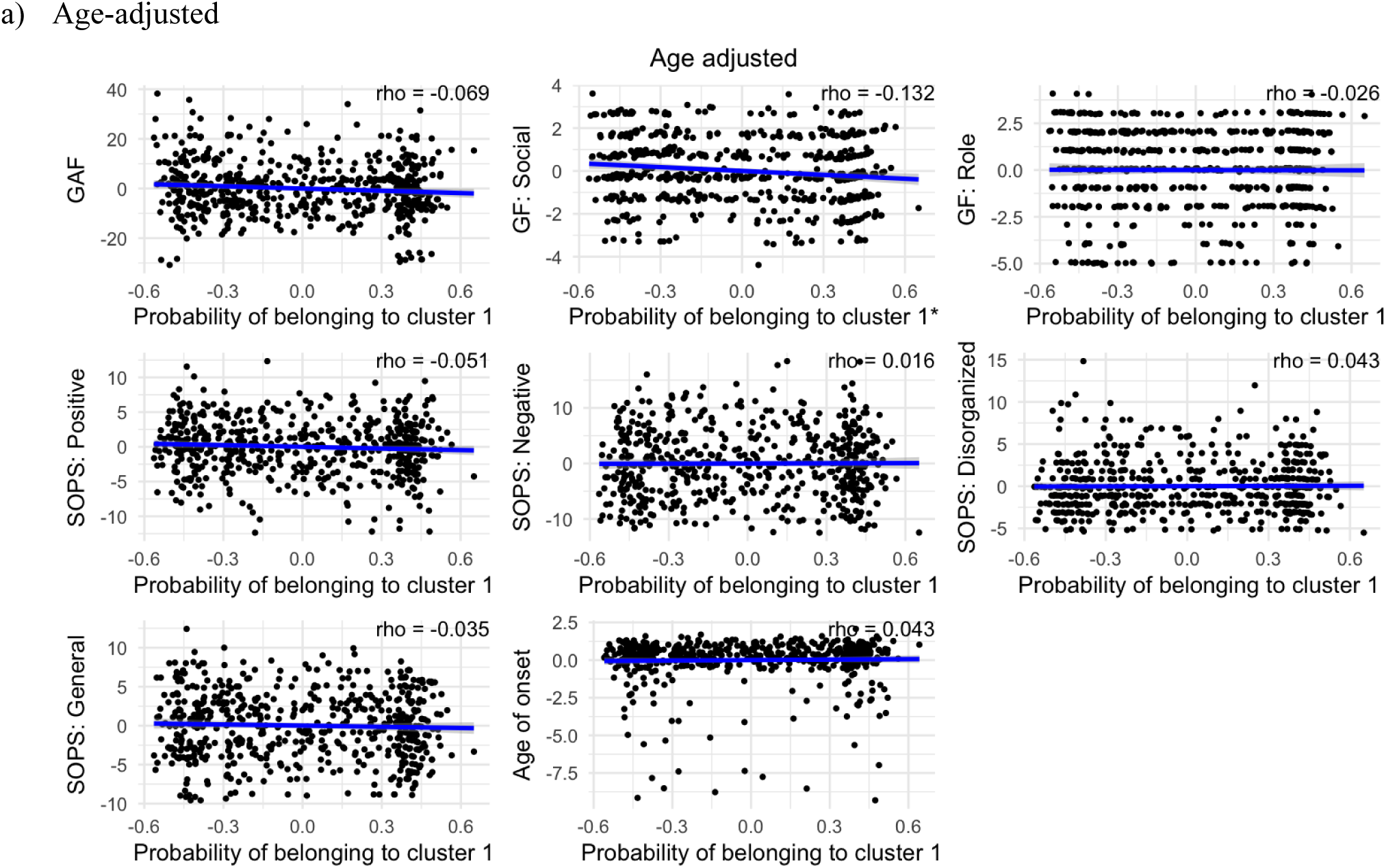
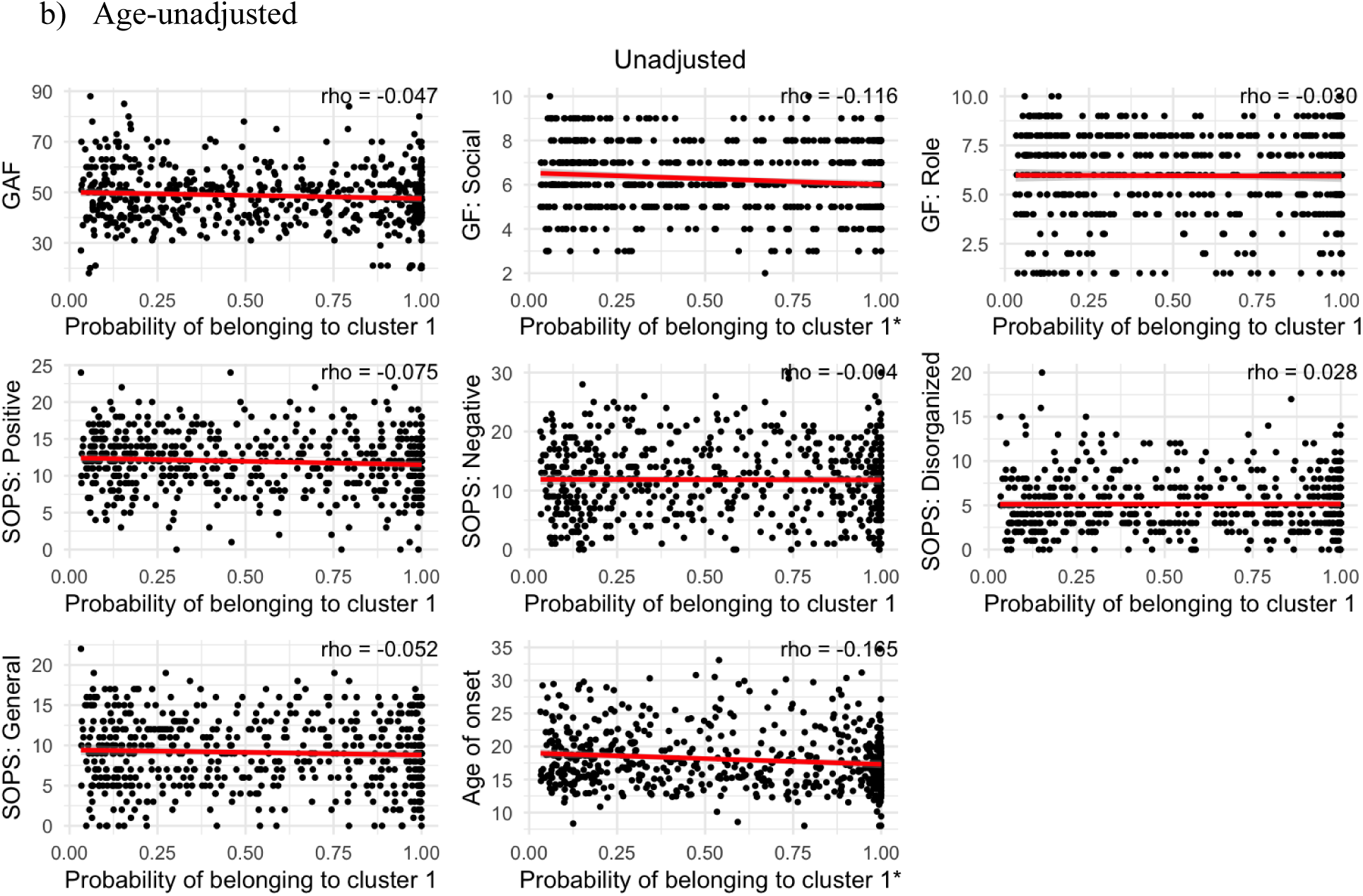
a,b: Age-adjusted and unadjusted association between clinical and functional outcomes with posterior probabilities in CHR participants from the NAPLS2. *CHR: Clinical high risk for psychosis; GAF: Global Assessment of Functioning; GF: Social: Global Social Functioning; GF: Role: Global Role Functioning; SOPS: Scale of Psychosis-Risk Symptoms; *: p<0.05; probabilities of a participant being in cluster 1 are posterior probabilities*

**Table 3:**
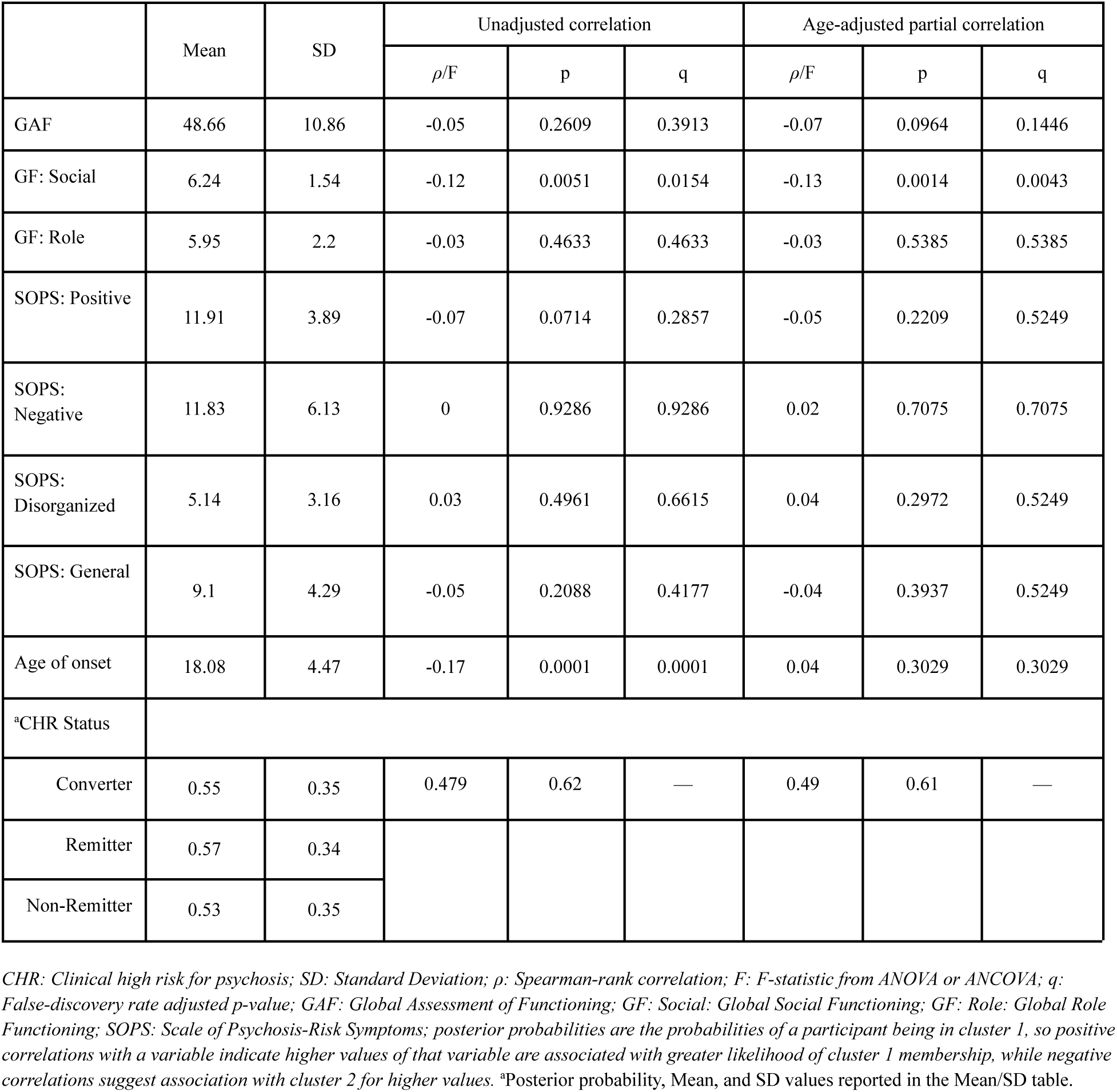
Age-adjusted and unadjusted association between clinical and functional outcomes with posterior probabilities in CHR participants from the NAPLS2.

|  | Mean | SD | Unadjusted correlation |  |  | Age-adjusted partial correlation |  |  |
| --- | --- | --- | --- | --- | --- | --- | --- | --- |
| | | | $\rho/F$ | p | q | $\rho/F$ | p | q |
| GAF | 48.66 | 10.86 | -0.05 | 0.2609 | 0.3913 | -0.07 | 0.0964 | 0.1446 |
| GF: Social | 6.24 | 1.54 | -0.12 | 0.0051 | 0.0154 | -0.13 | 0.0014 | 0.0043 |
| GF: Role | 5.95 | 2.2 | -0.03 | 0.4633 | 0.4633 | -0.03 | 0.5385 | 0.5385 |
| SOPS: Positive | 11.91 | 3.89 | -0.07 | 0.0714 | 0.2857 | -0.05 | 0.2209 | 0.5249 |
| SOPS: Negative | 11.83 | 6.13 | 0 | 0.9286 | 0.9286 | 0.02 | 0.7075 | 0.7075 |
| SOPS: Disorganized | 5.14 | 3.16 | 0.03 | 0.4961 | 0.6615 | 0.04 | 0.2972 | 0.5249 |
| SOPS: General | 9.1 | 4.29 | -0.05 | 0.2088 | 0.4177 | -0.04 | 0.3937 | 0.5249 |
| Age of onset | 18.08 | 4.47 | -0.17 | 0.0001 | 0.0001 | 0.04 | 0.3029 | 0.3029 |
| *CHR Status |  |  |  |  |  |  |  |  |
| Converter | 0.55 | 0.35 | 0.479 | 0.62 | — | 0.49 | 0.61 | — |
| Remitter | 0.57 | 0.34 |  |  |  |  |  |  |
| Non-Remitter | 0.53 | 0.35 |  |  |  |  |  |  |

CHR: Clinical high risk for psychosis; SD: Standard Deviation; $\rho$ : Spearman-rank correlation; F: F-statistic from ANOVA or ANCOVA; q: False-discovery rate adjusted p-value; GAF: Global Assessment of Functioning; GF: Social: Global Social Functioning; GF: Role: Global Role Functioning; SOPS: Scale of Psychosis-Risk Symptoms; posterior probabilities are the probabilities of a participant being in cluster 1, so positive correlations with a variable indicate higher values of that variable are associated with greater likelihood of cluster 1 membership, while negative correlations suggest association with cluster 2 for higher values. \*Posterior probability, Mean, and SD values reported in the Mean/SD table.

#### Conversion and remitter status

There was no significant association between clinical outcome status (converter, persistent CHR, or remitter) and the likelihood of belonging to cluster C1 versus C2 (Table 3).

### NAPLS-3

#### Demographics

Similar to the NAPLS-2 cohort, the NAPLS-3 participants who were more likely to be in C2 were older (p <0.001) and had more years of education (p <0.001) (Table S4). Both NAPLS-2 and NAPLS-3 had around a one-year average difference in age and education between the C2 and C1 clusters. Also, similar to NAPLS-2, NAPLS-3 had no statistically significant differences in sex or income between the clusters. CHR participants in NAPLS3 generally had higher cognition, smaller amplitudes in frontal and central MMN, N1, larger amplitudes in frontal and central AOD and VOD, and smaller amplitudes in parietal AOD and VOD (Table S5).

#### Cognition and EEG

NAPLS-3 participants had significantly higher cognition in C2, compared to C1, in most cognitive variables. The two clusters identified in NAPLS-3 were not different in WRAT, WASI IQ, and LNS (Table S6). The C1 cluster in NAPLS-3, relative to C2, had larger ERP amplitudes, including more negative MMN and N1(except for the central electrode sites), and more positive P3 across frontal, central, and parietal electrode sites (Figure S2a-b).

#### Clinical and Functional Outcomes

In the NAPLS-3 data, there were no significant associations between posterior probabilities of cluster C1 vs. C2 membership and clinical or functional outcomes (Figure S3a-b, Table S7). Sensitivity analyses can be found in Table S8.

NAPLS-2 data are available upon reasonable request, while NAPLS-3 data are available online at the National Institute of Mental Health Data Archive.

## Discussion

The present study utilized cognitive and EEG data from NAPLS-2 to identify subgroups within individuals at CHR and validate them using NAPLS-3. The clustering approach demonstrated weakly separable clusters. Thus, the study used *post hoc* probabilistic cluster membership, or posterior probabilities, rather than the initially planned discrete categorization. The results showed that CHR individuals who are more likely to be in C1 exhibited poorer cognitive performance across the board, as well as worse social functioning and an earlier age of symptom onset than C2. Interestingly, the C1 cluster, relative to C2, was associated with larger ERP component amplitudes, which were evident for MMN, auditory N1, and both auditory and visual target P3. NAPLS-3 results appeared to be largely similar to the results obtained from NAPLS 2.

Having poorer cognitive performance, worse functioning, and an earlier age of symptom onset within the same cluster aligns with a substantial body of literature across the psychosis spectrum, including evidence from NAPLS-2 that poorer cognition and worse function are associated with increased psychosis risk.^1,65–67^ On the other hand, in our sample, we see higher amplitudes of all three ERP components (MMN, auditory N1, auditory and visual P3) in the cluster having worse cognition and functioning. This might seem paradoxical, since the prior literature, including data published to date from NAPLS-2, shows lower amplitudes of these ERP components to be associated with increased psychosis risk.^24,68–76^ However, such results typically compare CHR with HC, not a comparison between CHR subgroups. In fact, studies comparing CHR subgroups, especially related to conversion, show higher P300 amplitude in more severe cases, or when autism is a comorbidity, for example.^77,78^ Thus, we should not assume that this neuropathophysiology develops linearly.

One possible interpretation is that these results reflect altered sensory gain regulation. When high gain is at hand, or when the brain increases the responsiveness of the auditory cortex, N1 would typically show a larger amplitude, and the opposite is true when we observe reduced responsiveness or lower gain.^79,80^ Moreover, MMN amplitude has been conceptualized within predictive coding frameworks as reflecting the precision weighting of auditory prediction errors.^81,82^ This could potentially lead to an enhanced bottom-up signal strength, which increases demand on executive filtering systems that are critical for working memory performance.^83^ Combined with the current study’s working memory findings, it is possible that overexcitation of the prefrontal cortex may reflect impaired working memory and cognitive flexibility.^84^ Thus, amplified early sensory and later attentional responses may reflect inefficient neural regulation that contributes to reduced cognitive efficiency, particularly in verbal working memory. This interpretation is consistent with evidence that sensory processing abnormalities and executive dysfunction are linked in psychosis-spectrum conditions. ^85^ Additionally, given the nature of MMN, it is more likely that the pathology is not limited to top-down processing but may reflect early sensory processing aberrations.^13^ Nonetheless, updating attentional resource allocation may not map directly onto specific cognitive domain severity assessed in our study, but it could be related to lower functioning. To this point, prior work does demonstrate that impairments in such neural domains are heterogeneous and can reflect state-like deficits as well as compensatory developmental processes rather than core markers of pathophysiology. ^78,86,87^

The good separability we saw when we clustered the cognitive variables alone in our previous work,^1^ and the poor separability seen when clustering cognition and EEG together, or EEG alone, highlights a broader methodological challenge in the field, such as the “kitchen sink” approach to clustering without explicitly acknowledging the developmental trajectory of the modalities used. Cognition and electrophysiology may not optimally combine into a single clustering solution because they might reflect overlapping and partially different developmental processes that diverge within the CHR at consecutive developmental time frames.

Notably, cognitive impairment in individuals at genetic high risk is evident early in development, preceding the emergence of attenuated psychotic symptoms by several years.^5,8,88,89^ In contrast, electrophysiological impairments in CHR, such as P300, appear to show more noticeable lower amplitude, relative to controls, closer to the age of symptom onset, and can be more vulnerable to maturational processes and illness progression or emerging circuit-level dysfunction.^90^ However, research investigating cognition in CHR longitudinally demonstrates stability.^3^ This may reflect the developmental timing differences between cognition and electrophysiological processes. It could also indicate that domain heterogeneity would require time to diverge, i.e., first establishing the core pathology and then later forming different levels, or clusters, of it.^91^

Given the separable clusters found in established psychosis when combining cognition and EEG, albeit using different EEG/ERP measures than those analyzed here, it is possible that, at the chronic stage of psychotic disorders, electrophysiological impairment has already diverged.^44,45^ This could also suggest that cognitive impairment may reflect a relatively stable vulnerability marker or an earlier trait-like change, while electrophysiological measures may be more sensitive to illness progression and circuit-level changes during the CHR phase or act as a later-emerging state-like change.

Our results are consistent with such an interpretation, given the observed partial overlap. Meaning that, at the CHR level, when one modality was less divergent than the other, rather than converging onto a unified subgroup structure, the modalities may have indexed distinct axes of heterogeneity that only partially intersected within individuals. When we evaluated the relationship between illness duration and uncertainty, we observed that the higher the illness duration, the more certain the model is about the subgroup the participant is placed in. This is consistent with the McGorry et al. psychosis staging framework. ^91^

Our findings suggest that a multimodal approach to clustering may not uniformly enhance subgroup delineation without theoretical guidance regarding developmental timing and the mechanisms of the modalities used. In CHR specifically, cognition can serve as a more stable measure of early risk and functional prognosis, whereas electrophysiology may be more suitable to track differentiation of illness categories, illness progression, and transition, and can play an important role as a primary biomarker in clinical trials given its lower heterogeneity at this developmental stage (Figure 4). Given these modalities’ complementary strengths and challenges, they can be highly synergistic in helping us understand the underlying mechanics of psychosis if used wisely.

**Figure 4:**
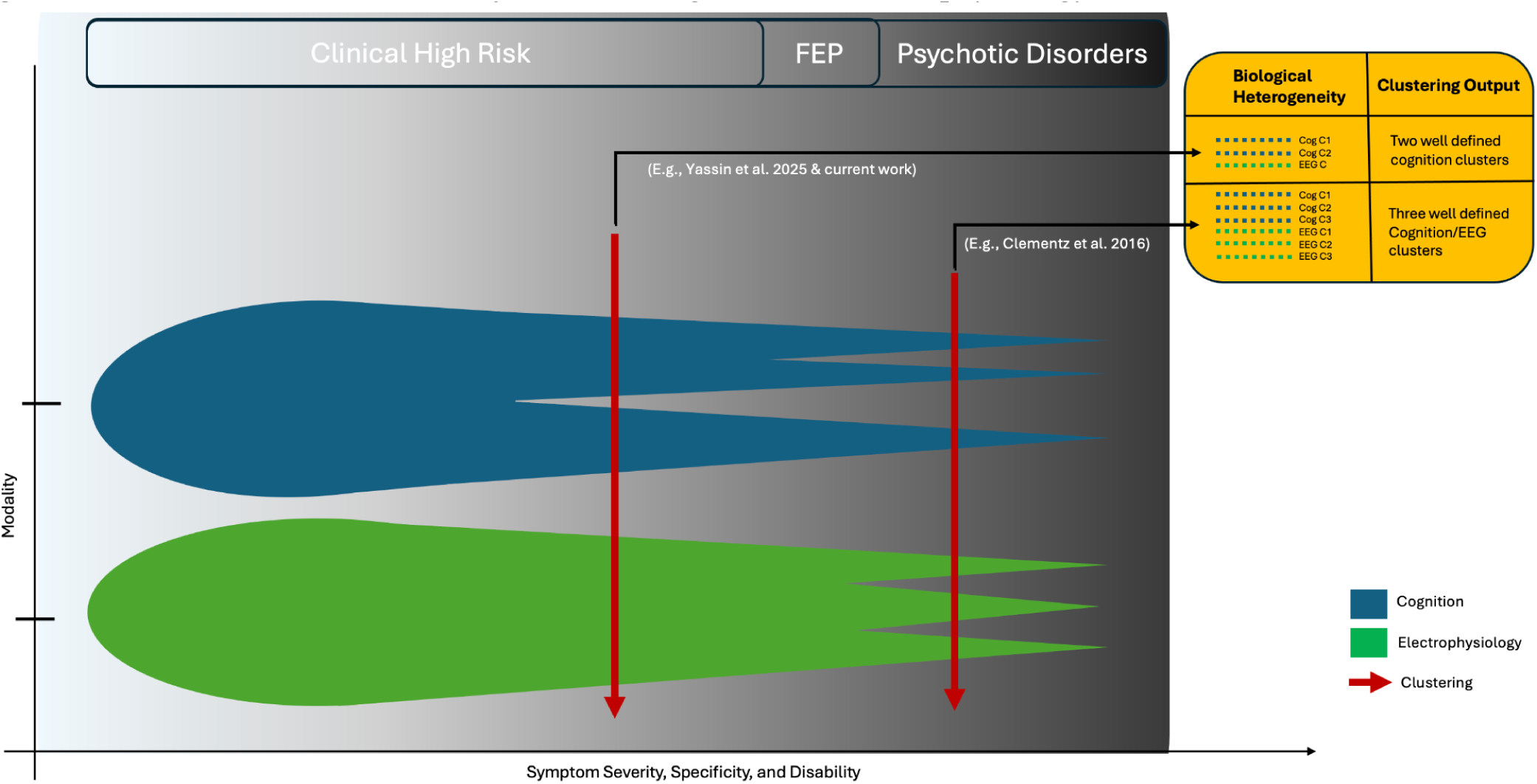
Schema of the theoretical trajectories of cognition and electrophysiology FEP: First Episode Psychosis; C: Cluster; Cog: Cognition; EEG: Electrophysiology *Supplementary materials*

## Limitations

This study has the following limitations: 1) Some of the electrophysiology measures in NAPLS 2 and 3 are not exactly the same. Nonetheless, they are similar in terms of the neurophysiological dimensions they inform. Despite that, observing similar results in both is reassuring. 2) The discussion section proposes that there is a developmental difference in terms of how different modalities cluster, and that future research should take this into account when performing clustering in CHR and in psychiatry as a whole.

The proposed hypothetical model across the lifespan needs to be tested in a longitudinal dataset that includes a much longer follow-up period, preferably evaluating genetically high-risk participants in their early stages of development. The current baseline data do not provide us with a definite answer to the proposed model; however, combined with previous evidence in CHR, FEP, and established psychosis disorders, the conclusion provides further support.

## Conclusion

Although combined cognitive and electrophysiology clustering yielded similar patterns across two large CHR cohorts, the weak separability suggests that multimodal clustering, if not developmentally grounded, may obscure meaningful substyping. Future work should prioritize longitudinal, modality-specific, and developmentally grounded approaches to parsing heterogeneity in psychosis risk; otherwise, if modalities such as those used in the current study were used uninformatively, we could be conflating early developmental cognitive risk markers with later-emerging neurophysiological alterations, resulting in clusters that are statistically identifiable but biologically intermingled, diffuse, and less meaningful.

## Data Availability

https://nda.nih.gov/

## Acknowledgements

We are thankful to the participants in this study. This work was supported by the National Institute of Mental Health (R21MH133001, WY; U01MH081984, JA; R01MH60720, U01MH081944 and K24MH76191, KSC; P50MH066286, CB; U01MH082004, DP; U01MH081988, EW; U01MH082022, SW/KSC; U01MH081928, WSS; U01MH081902, TC/CB; U01MH076989, DM; U01MH081857, BAC), made possible, first and foremost, by the participants in the study, the study team, and the taxpayers of the United States.

