## Supplemental Material for "Cognition and Electrophysiology Clustering in Clinical High Risk for Psychosis Delineates Distinct Dimensions of Heterogeneity: Implications for Multimodal Clustering"

### Supplementary material

**Table S1:** Comparison of NAPLS2 and NAPLS3 demographics between CHR and control participants.

| | NAPLS2 (N = 1044) | | | | | | NAPLS3 (N = 712) | | | | | |
|---|---|---|---|---|---|---|---|---|---|---|---|---|
| | Control (N=280) | | CHR (N=764) | | | | Control (N=84) | | CHR (N=628) | | | |
| | M or N | SD or % | M or N | SD or % | *t or* $X^2$ | p | M or N | SD or % | M or N | SD or % | *t or* $X^2$ | p |
| Age | 19.73 | 4.67 | 18.50 | 4.23 | 3.85 | <0.001 | 18.63 | 4.26 | 18.25 | 4.09 | 0.76 | 0.44 |
| Education (years) | 12.7 | 3.6 | 11.2 | 2.79 | 5.96 | <0.001 | 12.2 | 3.38 | 11.4 | 3.06 | 2.02 | <0.05 |
| Sex, male: female | 141 | 50.4% | 436 | 57.1% | 3.47 | 0.06 | 41 | 48.8% | 342 | 54.5% | 0.73 | 0.39 |
| Household Income | | | | | 12.49 | 0.05 | | | | | 7.99 | 0.239 |
| Less than 10,000 | 23 | 8.2% | 72 | 9.5% | | | 9 | 10.7% | 52 | 8.3% | | |
| 10,000 – 19,999 | 20 | 7.2% | 73 | 9.7% | | | 10 | 11.9% | 39 | 6.3% | | |
| 20,000 – 39,999 | 37 | 13.3% | 86 | 11.4% | | | 9 | 10.7% | 57 | 9.1% | | |
| 40,000 – 59,999 | 33 | 11.8% | 78 | 10.3% | | | 8 | 9.5% | 58 | 9.3% | | |
| 60,000 – 99,999 | 55 | 19.7% | 106 | 14.0% | | | 16 | 19.0% | 92 | 14.8% | | |
| 100,000 + | 68 | 24.4% | 170 | 22.5% | | | 18 | 21.4% | 171 | 27.4% | | |
| Don't know or refused | 43 | 15.4% | 171 | 22.6% | | | 14 | 16.7% | 154 | 24.7% | | |

*CHR: Clinical high risk for psychosis; M: Mean; SD: standard deviation; t: t-Test Statistic;* $X^2$*: Chi-square Statistic.*

**Table S2a-c:** Silhouette Scores

a) Clustering using electrophysiological features only in the NAPLS2.

| | Silhouette scores | | | | | | | | |
|---|---|---|---|---|---|---|---|---|---|
| Cluster Number | 2 | 3 | 4 | 5 | 6 | 7 | 8 | 9 | 10 |
| Diana | 0.35 | 0.294 | 0.289 | 0.253 | 0.235 | 0.23 | 0.228 | 0.213 | 0.201 |
| Agnes | 0.308 | 0.292 | 0.258 | 0.218 | 0.207 | 0.201 | 0.186 | 0.176 | 0.176 |
| K - means | 0.316 | 0.3 | 0.251 | 0.209 | 0.147 | 0.131 | 0.144 | 0.135 | 0.127 |

b) Clustering using electrophysiology and cognition features in the NAPLS2.

| | Silhouette scores | | | | | | | | |
|---|---|---|---|---|---|---|---|---|---|
| Cluster Number | 2 | 3 | 4 | 5 | 6 | 7 | 8 | 9 | 10 |
| Diana | 0.306 | 0.268 | 0.247 | 0.208 | 0.195 | 0.168 | 0.168 | 0.181 | 0.178 |
| Agnes | 0.248 | 0.231 | 0.164 | 0.143 | 0.142 | 0.138 | 0.156 | 0.156 | 0.156 |
| K - means | 0.292 | 0.27 | 0.209 | 0.173 | 0.2 | 0.166 | 0.161 | 0.129 | 0.123 |

c) Clustering using electrophysiology and cognition features in the NAPLS3.

| | Silhouette scores | | | | | | | | |
|---|---|---|---|---|---|---|---|---|---|
| Cluster Number | 2 | 3 | 4 | 5 | 6 | 7 | 8 | 9 | 10 |
| Diana | 0.401 | 0.203 | 0.206 | 0.112 | 0.112 | 0.112 | 0.109 | 0.109 | 0.106 |
| Agnes | 0.386 | 0.145 | 0.118 | 0.076 | 0.079 | 0.068 | 0.071 | 0.07 | 0.073 |
| K - means | 0.313 | 0.152 | 0.161 | 0.118 | 0.1 | 0.087 | 0.095 | 0.086 | 0.093 |

**Table S3:** Sensitivity analysis of clinical and functional outcome models comparing posterior probability estimates when including age or age and education as covariates while predicting outcomes from the NAPLS2 dataset.

| Outcome | *Comparison of beta estimates of posterior probabilities across models with age only, and age and education* | |
|---|---|---|
| | *Z* | *p* |
| GAF | -0.40 | 0.69 |
| GF: Social | -0.24 | 0.81 |
| GF: Role | -0.59 | 0.56 |
| SOPS: Positive | 0.00 | 1.00 |
| SOPS: Negative | 0.31 | 0.76 |
| SOPS: Disorganized | 0.00 | 1.00 |
| SOPS: General | 0.25 | 0.81 |
| Age of onset | -0.13 | 0.89 |

*This analysis compared the beta estimates of posterior probabilities between two regression models, the first with only age as a covariate predicting the outcome, and the second with age and education as covariates when predicting the outcome (e.g., GAF ~ Posterior Probability + Age + Education). The Z values statistical test statistic compares the degree of difference between the beta values, and the p-values are the significance value of that difference. GAF: Global Assessment of Functioning; GF: Social: Global Social Functioning; GF: Role: Global Role Functioning; SOPS: Scale of Psychosis-Risk Symptoms; Z: Wald test z-statistic; p: p-value of Wald test.*

| | C1 (N=122) | | C2 (N=356) | | *ρ/F* | *DF* | P |
|---|---|---|---|---|---|---|---|
| | M/N | SD/% | M/N | SD/% | | | |
| Age | 17.82 | 4.01 | 18.7 | 4.05 | -0.19 | – | <0.001*** |
| Education (years) | 11.16 | 3.04 | 11.85 | 3.01 | -0.20 | – | <0.001*** |
| Sex (male) | 66 | 54.10% | 194 | 54.49% | 0.00 | 1 | 0.98 |
| Household Income | | | | | 1.31 | 5 | 0.26 |
| Less than 10,000 | 100 | 86.21% | 272 | 81.19% | | | |
| 10,000 – 19,999 | 9 | 7.76% | 26 | 7.76% | | | |
| 20,000 – 39,999 | 2 | 1.72% | 24 | 7.16% | | | |
| 40,000 – 59,999 | 2 | 1.72% | 7 | 2.09% | | | |
| 60,000 – 99,999 | 2 | 1.72% | 1 | 0.30% | | | |
| 100,000 + | 1 | 0.86% | 5 | 1.49% | | | |

**Table S4:** CHR participant demographic data in the NAPLS3 by cluster membership.

*CHR: Clinical high risk for psychosis; C1: cluster 1; C2: cluster 2; M/N: Mean/ category count for participants in the given cluster; SD/%: Standard deviation/ category percentage for participants in the given cluster; ρ/F: Overall Spearman-rank correlation coefficient with respect to posterior probability/ ANOVA F-statistics with respect to posterior probability; DF: degrees of freedom; P: p-value.*

**Table S5:** Cognition and Electrophysiology in NAPLS2 and NAPLS3 CHR participants

| Domains | | NAPLS2 | | NAPLS3 | | | |
|---|---|---|---|---|---|---|---|
| | | Mean | SD | Mean | SD | T | p |
| WRAT | | 50.55 | 6.74 | 60.22 | 6.95 | -22.84 | 0.0000 |
| WASI: Vocab | | 38.62 | 9.07 | 38.84 | 6.29 | -0.47 | 0.6353 |
| WASI IQ | | 99.12 | 14.45 | 106.46 | 16.11 | -7.72 | 0.0000 |
| CPT: QA | | 91.55 | 7.30 | 95.84 | 6.63 | -9.95 | 0.0000 |
| CPT: Q3A | | 70.97 | 14.35 | 81.84 | 13.33 | -12.66 | 0.0000 |
| CPT: Interference | | 63.74 | 19.97 | 76.95 | 16.53 | -11.70 | 0.0000 |
| BACS | | 43.82 | 12.49 | 55.05 | 13.60 | -13.88 | 0.0000 |
| HVLT | | 22.38 | 4.77 | 26.51 | 5.23 | -13.32 | 0.0000 |
| LNS | | 11.99 | 3.41 | 14.89 | 3.59 | -13.39 | 0.0000 |
| MMN | F3 | -3.05 | 2.15 | -1.01 | 1.00 | -20.39 | 0.0000 |
| | Fz | -3.12 | 2.30 | -1.50 | 1.14 | -14.92 | 0.0000 |
| | F4 | -3.42 | 2.21 | -1.30 | 1.10 | -20.33 | 0.0000 |
| N100 | F3stanAOD | -2.92 | 2.30 | -1.21 | 1.14 | -15.68 | 0.0000 |
| | FzstanAOD | -3.78 | 2.54 | -1.93 | 1.35 | -15.06 | 0.0000 |
| | F4stanAOD | -2.95 | 2.36 | -1.53 | 1.26 | -12.39 | 0.0000 |
| | F3targAOD | -7.10 | 3.74 | -2.07 | 1.73 | -28.63 | 0.0000 |
| | FztargAOD | -8.13 | 3.77 | -2.85 | 1.83 | -29.52 | 0.0000 |
| | F4targAOD | -7.56 | 3.70 | -2.52 | 1.84 | -28.51 | 0.0000 |
| P300 | F3targAOD | -0.55 | 5.72 | -3.16 | 2.27 | 10.08 | 0.0000 |
| | FztargAOD | 0.44 | 6.27 | -2.96 | 2.74 | 11.77 | 0.0000 |
| | F4targAOD | -0.42 | 5.59 | -2.10 | 2.45 | 6.51 | 0.0000 |
| | FztargVOD | 4.08 | 12.30 | -3.65 | 3.12 | 14.61 | 0.0000 |
| | F4targVOD | 4.79 | 11.52 | -2.59 | 2.78 | 14.96 | 0.0000 |
| | F3targVOD | 3.52 | 11.10 | -3.76 | 2.80 | 15.25 | 0.0000 |
| MMN | C3 | -2.72 | 2.13 | -0.76 | 0.72 | -20.87 | 0.0000 |
| | Cz | -2.58 | 2.28 | -1.11 | 0.96 | -14.18 | 0.0000 |
| | C4 | -3.12 | 2.14 | -0.80 | 0.82 | -24.08 | 0.0000 |
| N100 | C3stanAOD | -2.53 | 2.24 | -1.26 | 1.06 | -12.11 | 0.0000 |
| | CzstanAOD | -2.62 | 2.38 | -2.12 | 1.31 | -4.25 | 0.0000 |
| | C4stanAOD | -2.61 | 2.10 | -1.47 | 1.10 | -11.23 | 0.0000 |
| | C3targAOD | -6.59 | 3.62 | -2.04 | 1.50 | -27.39 | 0.0000 |
| | CztargAOD | -5.96 | 3.67 | -2.79 | 1.72 | -18.37 | 0.0000 |
| | C4targAOD | -6.76 | 3.33 | -2.28 | 1.56 | -28.60 | 0.0000 |
| P300 | C3targAOD | 4.92 | 5.61 | 0.79 | 2.08 | 16.40 | 0.0000 |
| | CztargAOD | 7.84 | 6.64 | 2.33 | 2.80 | 18.12 | 0.0000 |
| | C4targAOD | 4.90 | 5.40 | 1.87 | 1.82 | 12.69 | 0.0000 |
| | CztargVOD | 13.41 | 13.14 | 2.90 | 3.01 | 18.74 | 0.0000 |
| | C3targVOD | 9.34 | 12.21 | 1.06 | 2.20 | 16.06 | 0.0000 |
| | C4targVOD | 9.53 | 11.78 | 2.29 | 2.05 | 14.61 | 0.0000 |
| | P3targAOD | 8.65 | 5.67 | 3.55 | 2.44 | 19.52 | 0.0000 |
| | P3targVOD | 12.91 | 12.76 | 4.59 | 2.87 | 15.29 | 0.0000 |
| | P4targAOD | 8.73 | 5.45 | 3.18 | 2.20 | 22.40 | 0.0000 |
| | P4targVOD | 12.30 | 12.33 | 4.49 | 2.61 | 14.91 | 0.0000 |
| | PztargAOD | 11.85 | 6.38 | 5.17 | 2.89 | 22.52 | 0.0000 |
| | PztargVOD | 18.08 | 13.44 | 7.31 | 3.38 | 18.65 | 0.0000 |
| | VODp3Latency | 444.31 | 46.99 | 442.33 | 59.67 | 0.59 | 0.5586 |
| | AODp3Latency | 316.38 | 33.70 | 321.50 | 45.57 | -2.00 | 0.0455 |

*WRAT: Wide Range Achievement Test-Four Reading subtest; WASI Vocab: Wechsler Abbreviated Scale for Intelligence-2 Vocabulary; WASI IQ: Weschler Abbreviated Scale for Intelligence - 2: Intelligence Quotient; CPT: Auditory Working Memory Continuous Performance Test; QA: Vigilance; Q3A: Working Memory Load/No Interference; BACS: Brief Assessment of Cognition in Schizophrenia – Symbol Coding; HVLT: Hopkins Verbal Learning Test-Revised; LNS: Letter-Number-Span; MMN: Mismatch negativity; AOD: Auditory Oddball task; VOD: Visual*

*Oddball Task; N1 (Or N100): Event related potential Negative peak around 100 milliseconds; P3 (Or P300): Event related potential Positive peak around 300 milliseconds; Targ: Target; Stan: Standard.*

**Table S6:** Correlations between cognitive and electrophysiology domains with posterior probabilities in CHR participants from the NAPLS3.

| Domains | | Mean | SD | | |
|---|---|---|---|---|---|
| | | | | $\rho$ | p |
| Cognition | WRAT | 60.22 | 6.95 | -0.09 | 0.0512 |
| | WASI: Vocab | 38.84 | 6.29 | -0.11 | 0.0141 |
| | WASI IQ | 106.46 | 16.11 | -0.07 | 0.1520 |
| | CPT: QA | 95.84 | 6.63 | -0.16 | 0.0007 |
| | CPT: Q3A | 81.84 | 13.33 | -0.15 | 0.0017 |
| | CPT: Interference | 76.95 | 16.53 | -0.18 | 0.0001 |
| | BACS | 55.05 | 13.6 | -0.12 | 0.0078 |
| | HVLT | 26.51 | 5.23 | -0.11 | 0.0135 |
| | LNS | 14.89 | 3.59 | -0.08 | 0.0884 |
| Frontal - MMN | MMNampF3 | -1.01 | 1 | -0.12 | 0.0070 |
| | MMNampFz | -1.5 | 1.14 | -0.1 | 0.0223 |
| | MMNampFz4 | -1.3 | 1.1 | -0.14 | 0.0016 |
| Frontal - N100 | F3stanAODn1Amp | -1.21 | 1.14 | -0.09 | 0.0602 |
| | FzstanAODn1Amp | -1.93 | 1.35 | -0.17 | 0.0004 |
| | F4stanAODn1Amp | -1.53 | 1.26 | -0.16 | 0.0005 |
| | F3targAODn1Amp | -2.07 | 1.73 | -0.16 | 0.0006 |
| | FztargAODn1Amp | -2.85 | 1.83 | -0.24 | 0.0000 |
| | F4targAODn1Amp | -2.52 | 1.84 | -0.23 | 0.0000 |
| Frontal - AOD | F3targAODAmp | -3.16 | 2.27 | -0.12 | 0.0113 |
| | FztargAODAmp | -2.96 | 2.74 | -0.07 | 0.1206 |
| | F4targAODAmp | -2.1 | 2.45 | -0.07 | 0.1508 |
| Frontal - VOD | FztargVODAmp | -3.65 | 3.12 | -0.12 | 0.0115 |
| | F4targVODAmp | -2.59 | 2.78 | -0.07 | 0.1452 |
| | F3targVODAmp | -3.76 | 2.8 | -0.06 | 0.1998 |
| Central - MMN | MMNampC3 | -0.76 | 0.72 | 0.01 | 0.7613 |
| | MMNampCz | -1.11 | 0.96 | 0.04 | 0.4445 |
| | MMNampC4 | -0.8 | 0.82 | -0.07 | 0.1531 |
| Central N100 | FC3stanAODn1Amp | -1.26 | 1.06 | -0.09 | 0.0512 |
| | FCzstanAODn1Amp | -2.12 | 1.31 | -0.17 | 0.0003 |
| | FC4stanAODn1Amp | -1.47 | 1.1 | -0.18 | 0.0001 |
| | FC3targAODn1Amp | -2.04 | 1.5 | -0.15 | 0.0022 |

| | | | | | |
|---|---|---|---|---|---|
| | FCztargAODn1Amp | -2.79 | 1.72 | -0.2 | 0.0000 |
| | FC4targAODn1Amp | -2.28 | 1.56 | -0.24 | 0.0000 |
| Central AOD | C3targAODAmp | 0.79 | 2.08 | 0.09 | 0.0527 |
| | CztargAODAmp | 2.33 | 2.8 | 0.11 | 0.0155 |
| | C4targAODAmp | 1.87 | 1.82 | 0.07 | 0.1292 |
| Central VOD | CztargVODAmp | 2.9 | 3.01 | 0.03 | 0.5379 |
| | C3targVODAmp | 1.06 | 2.2 | 0.05 | 0.3213 |
| | C4targVODAmp | 2.29 | 2.05 | -0.02 | 0.5968 |
| Parietal AOD | P3targAODAmp | 3.55 | 2.44 | 0.1 | 0.0278 |
| | PztargAODAmp | 5.17 | 2.89 | 0.1 | 0.0277 |
| | P4targAODAmp | 3.18 | 2.2 | 0.1 | 0.0410 |
| Parietal VOD | P4targVODAmp | 4.49 | 2.61 | 0.01 | 0.7845 |
| | P3targVODAmp | 4.59 | 2.87 | 0.09 | 0.0428 |
| | PztargVODAmp | 7.31 | 3.38 | 0.07 | 0.1215 |
| VODp3Latency | | 442.33 | 59.67 | 0.03 | 0.4599 |
| AODp3Latency | | 321.5 | 45.57 | -0.02 | 0.6661 |

*CHR: Clinical high risk for psychosis; SD: Standard deviation; ρ: Spearman-rank correlation; p: p-value of correlation test; WRAT: Wide Range Achievement Test-Four Reading subtest; WASI Vocab: Wechsler Abbreviated Scale for Intelligence-2 Vocabulary; WASI IQ: Weschler Abbreviated Scale for Intelligence - 2: Intelligence Quotient; CPT: Auditory Working Memory Continuous Performance Test; QA: Vigilance; Q3A: Working Memory Load/No Interference; BACS: Brief Assessment of Cognition in Schizophrenia – Symbol Coding; HVLT: Hopkins Verbal Learning Test-Revised; LNS: Letter-Number-Span; MMN: Mismatch negativity; AOD: Auditory Oddball task; VOD: Visual Oddball Task; N1 (Or N100): Event related potential Negative peak around 100 milliseconds; P3 (Or P300): Event related potential Positive peak around 300 milliseconds; Targ: Target; Stan: Standard; Amp: Amplitude.*

**Figure S1.** Relationship between illness duration up to 6 years and uncertainty of belonging to a specific cluster using Pearson correlation.

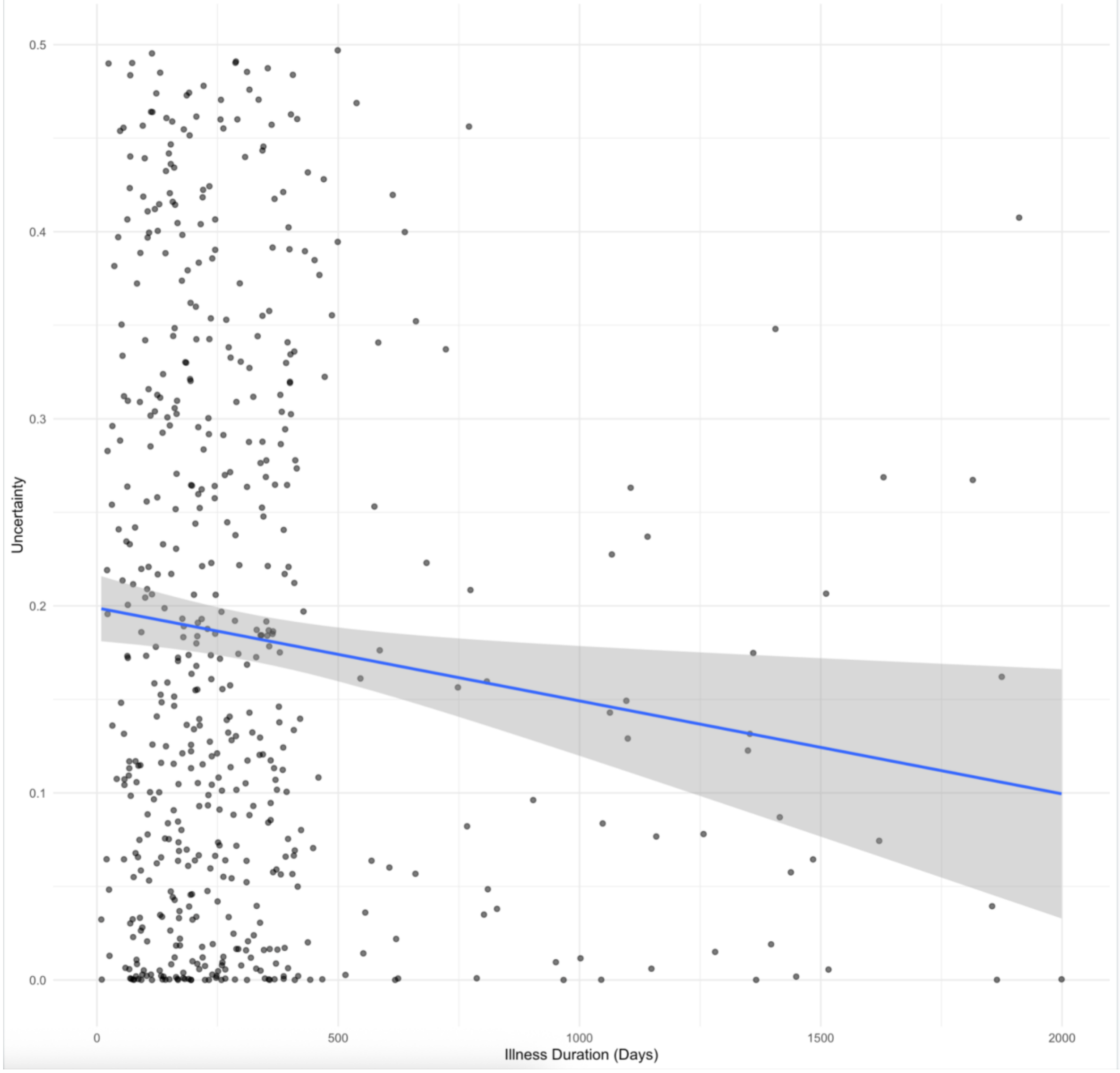


*Note: Pearson correlation r: -0.1064, p-value: 0.0124364*

**Figure S2a, b:** Correlations between cognitive/EEG variables with posterior probabilities in CHR participants from NAPLS3.

a) Cognition

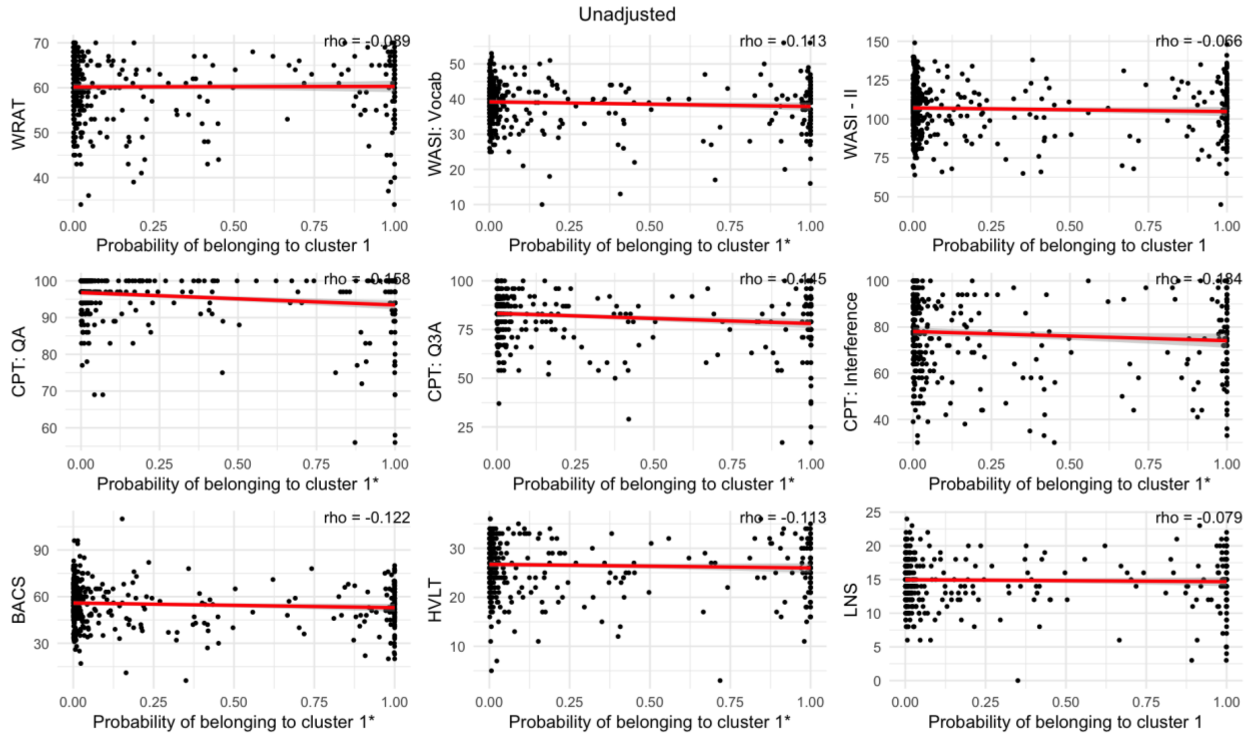

b) EEG

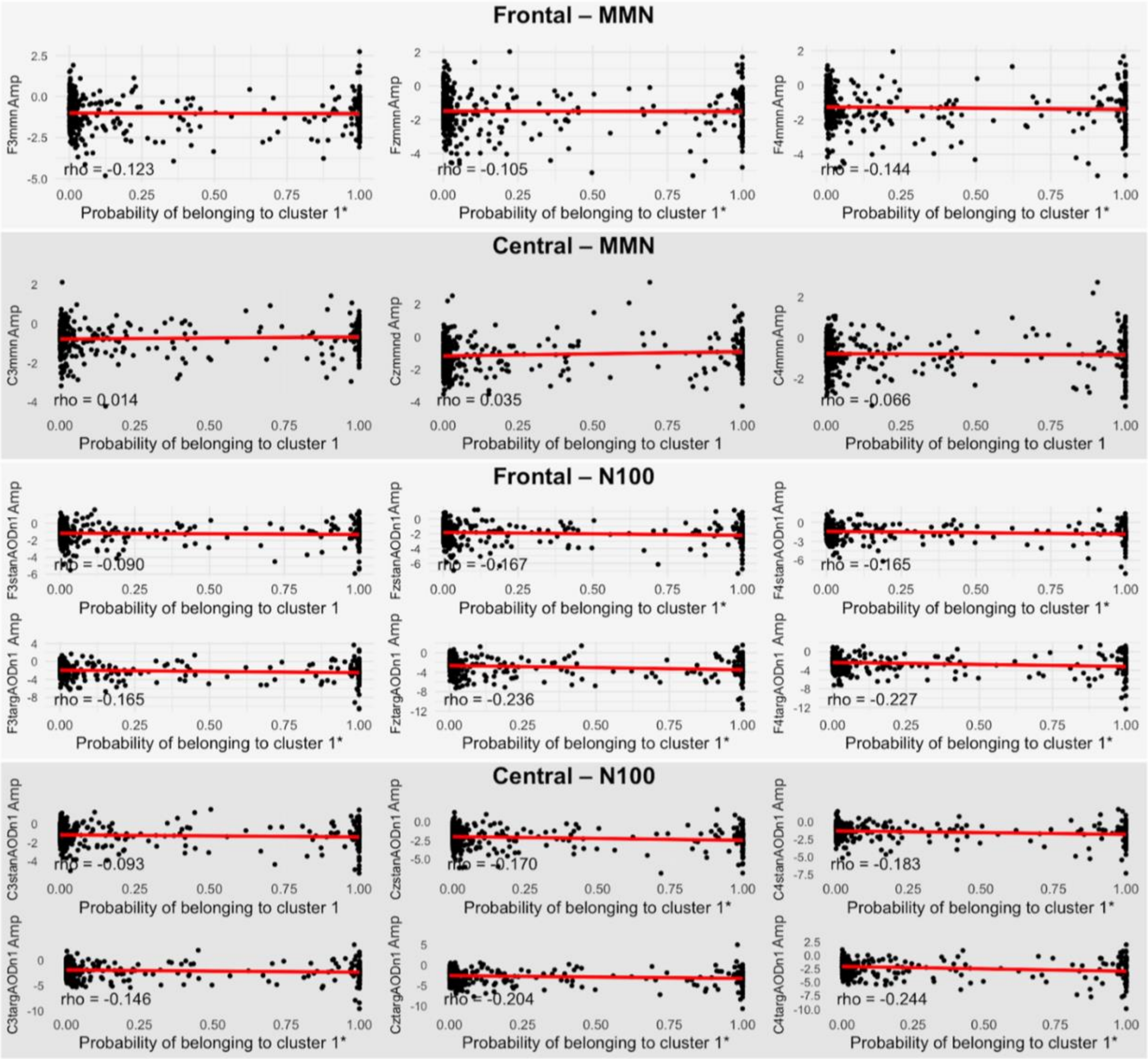
Frontal – MMN
F3mmn Amp
FzmmnAmp
F4mmn Amp
rho = -0.123
rho = -0.105
rho = -0.144
Probability of belonging to cluster 1*
Central – MMN
C3mmn Amp
Czmmnd Amp
C4mmn Amp
rho = 0.014
rho = 0.035
rho = -0.066
Probability of belonging to cluster 1
Frontal – N100
F3stanAODn1 Amp
FzstanAODn1 Amp
F4stanAODn1 Amp
rho = -0.090
rho = -0.167
rho = -0.165
F3targAODn1 Amp
FztargAODn1 Amp
F4targAODn1 Amp
rho = -0.165
rho = -0.236
rho = -0.227
Central – N100
C3stanAODn1 Amp
CzstanAODn1 Amp
C4stanAODn1 Amp
rho = -0.093
rho = -0.170
rho = -0.183
C3targAODn1 Amp
CztargAODn1 Amp
C4targAODn1 Amp
rho = -0.146
rho = -0.204
rho = -0.244

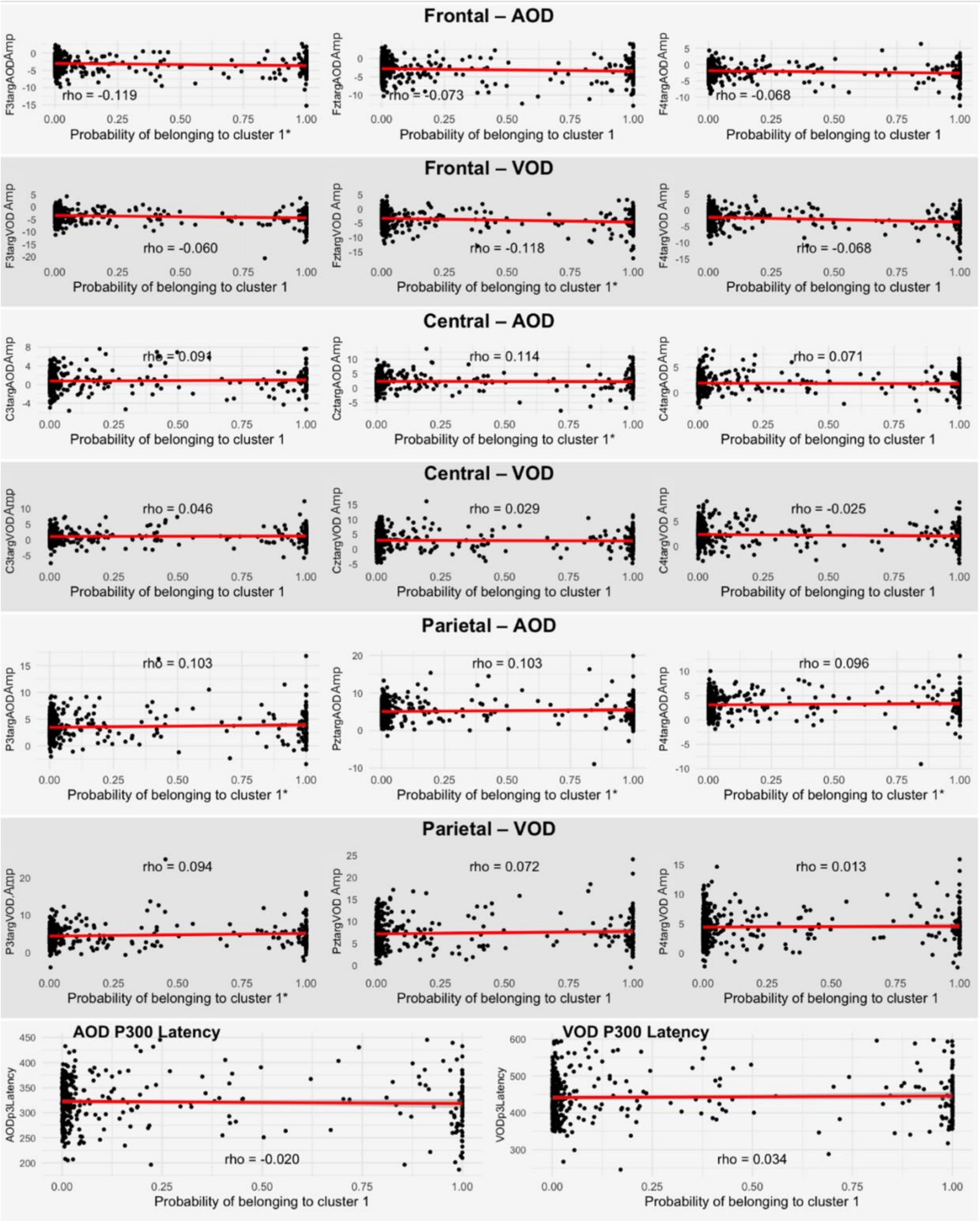


*CHR: Clinical high risk for psychosis; WRAT: Wide Range Achievement Test-Four Reading subtest; WASI Vocab: Wechsler Abbreviated Scale for Intelligence-2 Vocabulary; WASI - II: Weschler Abbreviated Scale for Intelligence - 2: Intelligence Quotient; CPT: Auditory Working Memory Continuous Performance Test; QA: Vigilance; Q3A: Working Memory Load/No Interference; BACS: Brief Assessment of Cognition in Schizophrenia – Symbol Coding; HVLT: Hopkins Verbal Learning Test-Revised; LNS: Letter-Number-Span; MMN: Mismatch negativity; AOD: Auditory Oddball task; VOD: Visual Oddball Task; N1 (Or N100): Event related potential; Negative peak around 100 milliseconds; P3 (Or P300): Event related potential Positive peak around 300 milliseconds; Targ: Target; Stan: Standard; Amp: Amplitude; *: p<0.05.*

**Table S7:** Age-adjusted and unadjusted correlations between clinical and functional outcomes with posterior probabilities in CHR participants from the NAPLS3 dataset.

| Outcome | Mean/N | SD/% | Not adjusted for age | | | Adjusted for age | | |
|---|---|---|---|---|---|---|---|---|
| | | | ρ/ OR (CI) | p | q | ρ/ OR (CI) | p | q |
| GAF | 51.11 | 12.41 | 0.03 | 0.5578 | 0.5578 | 0.04 | 0.3767 | 0.4674 |
| GF: Social | 6.44 | 1.51 | 0.03 | 0.4631 | 0.5578 | 0.03 | 0.4674 | 0.4674 |
| GF: Role | 6.37 | 2.22 | -0.06 | 0.2266 | 0.5578 | -0.08 | 0.0909 | 0.2727 |
| SOPS: Positive | 13.01 | 3.36 | -0.06 | 0.2228 | 0.6498 | -0.07 | 0.1311 | 0.5246 |
| SOPS: Negative | 12.18 | 6.38 | 0.03 | 0.4874 | 0.6498 | 0.04 | 0.4362 | 0.6851 |
| SOPS: Disorganized | 5.16 | 3.24 | 0.03 | 0.455 | 0.6498 | 0.03 | 0.5138 | 0.6851 |
| SOPS: General | 9.62 | 4.21 | 0.01 | 0.9088 | 0.9088 | 0 | 0.9964 | 0.9964 |
| Age of onset | 20.19 | 4.40 | -0.06 | 0.6772 | 0.6772 | 0.11 | 0.4918 | 0.4918 |
| Converter Status (converter vs. non-converter) | 44 | 9.21% | 1.74 (0.84 - 3.49) | 0.124 | – | 1.80 (0.87 - 3.64) | 0.104 | – |

*CHR: Clinical high risk for psychosis; N: category count for participants in the given cluster; SD/%: Standard deviation/ category percentage for participants in the given cluster; ρ: correlation; OR: Odd's ratio from logistic regression; CI: Confidence Intervals from logistic regression; p: p-value from correlation test or regression; q: False-discovery rate adjusted p-value; GAF: Global Assessment of Functioning; GF: Social: Global Social Functioning; GF: Role: Global Role Functioning; SOPS: Scale of Psychosis-Risk Symptoms; Data summary and analysis only accounted for CHR participants. Logistic regression was conducted for converter status, given that there were only two outcomes identified in NAPLS3 thus far (converter vs. non-converter).*

**Figure S3a,b:** Scatterplots of age-adjusted and unadjusted correlations between probabilities of being assigned cluster 1 with clinical and functional outcomes of CHR participants within the NAPLS3 cohort.

a) Unadjusted clinical and functional outcomes plotted against the probability of belonging to cluster 1.

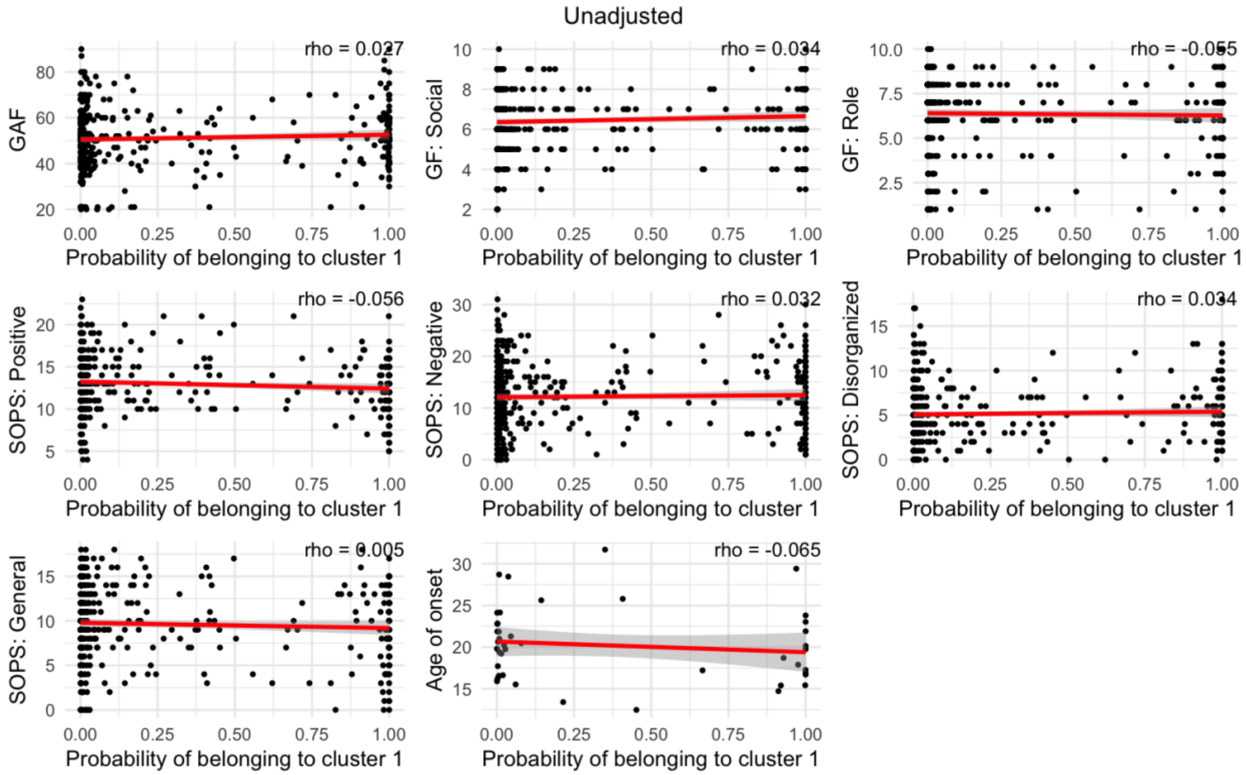

b) Age-adjusted residuals of clinical and functional outcomes plotted against age-adjusted residuals of cluster 1 probability

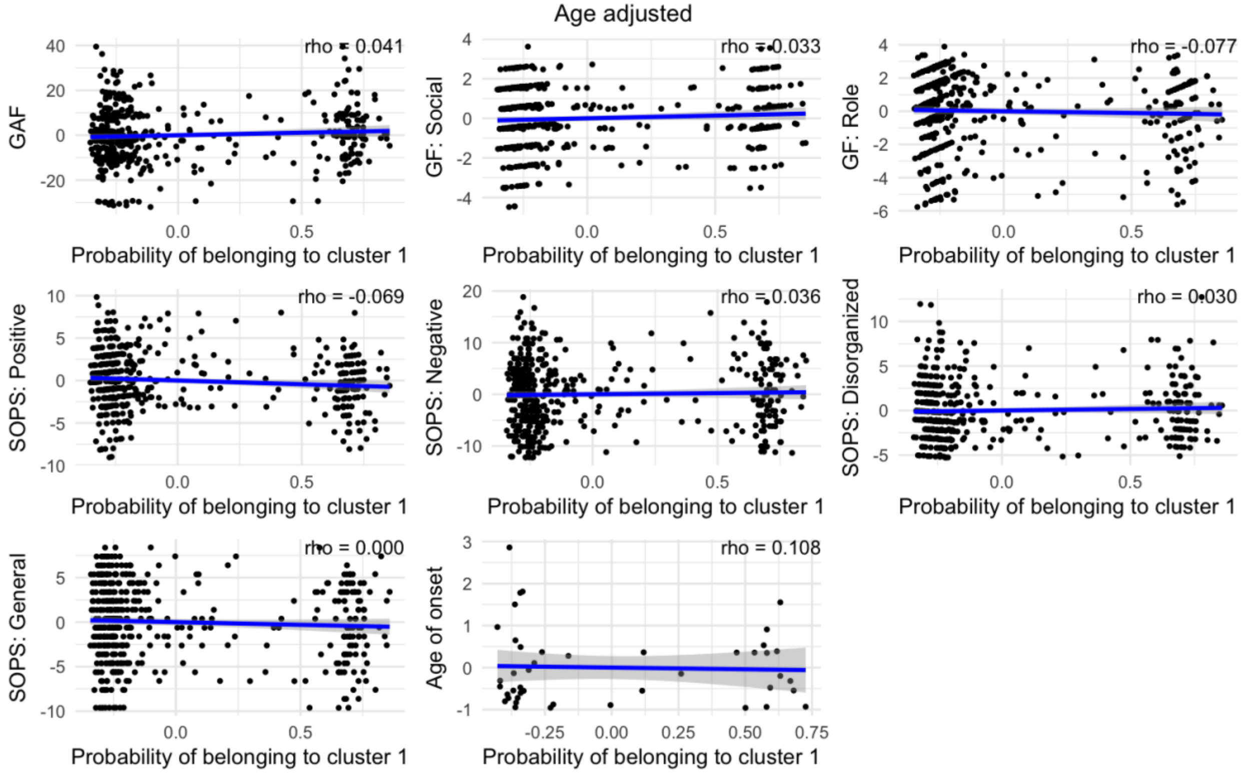


*CHR: Clinical high risk for psychosis; GAF: Global Assessment of Functioning; GF: Social: Global Social Functioning; GF: Role: Global Role Functioning; SOPS: Scale of Psychosis-Risk Symptoms; *: p<0.05.*

**Table S8:** Sensitivity analysis of clinical and functional outcome models comparing posterior probability estimates when including age or age and education as covariates while predicting outcomes from the NAPLS3 dataset.

| Outcome | *Comparison of beta estimates of posterior probabilities across models with age and age and education* | |
|---|---|---|
| | *Z* | *p* |
| GAF | -0.23 | 0.81 |
| GF: Social | -0.32 | 0.75 |
| GF: Role | -0.33 | 0.74 |
| SOPS: Positive | 0.12 | 0.91 |
| SOPS: Negative | 0.23 | 0.82 |
| SOPS: Disorganized | 0.31 | 0.76 |
| SOPS: General | 0.17 | 0.87 |
| Age of onset | -0.03 | 0.98 |

*This analysis compared the beta estimates of posterior probabilities between two regression models, the first with only age as a covariate predicting the outcome, and the second with age and education as covariates when predicting the outcome (e.g., GAF ~ Posterior Probability + Age + Education). The Z values statistical test statistic compares the degree of difference between the beta values, and the p-values are the significance value of that difference. GAF: Global Assessment of Functioning; GF: Social: Global Social Functioning; GF: Role: Global Role Functioning; SOPS: Scale of Psychosis-Risk Symptoms. Z: Wald test z-statistic; p: p-value of Wald test.*
